# A dynamic ensemble model for short-term forecasting in pandemic situations

**DOI:** 10.1101/2024.03.08.24303963

**Authors:** Jonas Botz, Diego Valderrama, Jannis Guski, Holger Fröhlich

## Abstract

During the COVID-19 pandemic, many hospitals reached their capacity limits and could no longer guarantee treatment of all patients. At the same time, governments endeavored to take sensible measures to stop the spread of the virus while at the same time trying to keep the economy afloat. Many models extrapolating confirmed cases and hospitalization rate over short periods of time have been proposed, including several ones coming from the field of machine learning. However, the highly dynamic nature of the pandemic with rapidly introduced interventions and new circulating variants imposed non-trivial challenges for the generalizability of such models.

In the context of this paper, we propose the use of ensemble models, which are allowed to change in their composition or weighting of base models over time and can thus adapt to highly dynamic pandemic or epidemic situations. In that regard, we also explored the use of secondary metadata - Google searches - to inform the ensemble model. We tested our approach using surveillance data from COVID-19, Influenza, and hospital syndromic surveillance of severe acute respiratory infections (SARI). In general, we found ensembles to be more robust than the individual models. Altogether we see our work as a contribution to enhance the preparedness for future pandemic situations.

## 1. Introduction

In late 2019 a novel coronavirus SARS-CoV-2 emerged [1]. This not only gave rise to the COVID-19 pandemic but also affected every aspect of human life, from an economic downturn, and disruption in education and social interactions to severe health implications including millions of deaths [2–4]. Early on, governments struggled to find a balance between containing the spread of the virus and maintaining as much economy, social interactions, and educational services as possible. Important indicators for decision-making were the number of confirmed cases and the hospitalization rate. During that time many models were developed for short-term forecasting of the number of incident cases and hospitalizations, respectively [5], modeling strategies in this field include mechanistic, machine learning, and hybrid modeling strategies [5]. All these models learn patterns from historical data to make forecasts, i.e. there is the implicit assumption of a stationary dynamical process. However, the highly dynamic nature of the pandemic with the rapid introduction of non-pharmaceutical interventions, new vaccines, and new circulating virus variants contradicted this assumption and thus imposed non-trivial challenges for the generalizability of all forecasting models over longer periods of time, regardless of the chosen modeling strategy.

Since each modeling technique unavoidably comes along with its own assumptions and limitations, ensemble models have been proposed for forecasting the spread of infectious diseases like Influenza [6–8] or Ebola [9] and later for COVID-19 [10,11]. In principle, ensemble models can be understood as a collection of rather simplistic base models, which all produce an output based on each model’s assumption plus an algorithm or meta-model that combines them into one ensemble output. The advantage of such an ensemble approach is that the bias of the individual models is reduced, making the final output more robust [12]. In the literature, such ensemble methods often use the mean (e.g., [11]) or median (e.g., [10]) of the base model outputs. However, pandemics like COVID-19 are dynamic, there are times when the number of cases barely changes, there is exponential growth and decay, and there are turning points of waves, which can all depend on external factors like interventions [13,14], people’s behavior [15], seasonality [16,17], or variants of concern [18,19].

To capture these dynamics and to be better prepared for future pandemics we here propose an ensemble modeling approach that is dynamically adjusted, to either select the right model at the right time or to weigh the models’ predictions according to the current situation by using a meta-model. As base models, we implemented a linear regression, ARIMA [20], XGBoost [21], Random Forest [22], and an LSTM [23] model. We then evaluated the performance of each base model and compared this to baseline ensemble methods. In the next step, we implemented a multi-layer perceptron (MLP) with softmax heads as a meta-model. The base models’ forecasts and performances were used as input for the meta-model which was trained in either one of two ways: 1. select one of the models (selection), 2. combine the model’s predictions into one prediction (stacking). In addition, we tested whether the inclusion of metadata coming from Google Trends could inform the meta-model to make better decisions.

## 2. Materials and Methods

### 2.1 Surveillance Data

We incorporate six different datasets related to COVID-19: the daily number of incident cases, hospital admissions (hospitalization), and deaths for Germany and France, respectively. Additionally, we evaluated the models on weekly Influenza cases and weekly hospital admissions related to severe acute respiratory infections (SARI) in Germany. While the weekly data is only available on a country level, the daily data is on a regional level (16 Bundesländer in Germany and 13 Régions in France, excluding overseas regions). Moreover, we also included the country-level data for the daily data. While in the SARI and Influenza datasets, the hospitalized and incident cases are provided, normalized to 100 thousand people (incidence), in the other datasets we worked with absolute numbers. An overview of the used surveillance data can be found in Table 1. The German surveillance data were received from the Robert Koch Institute (RKI) (https://github.com/robert-koch-institut) and the French surveillance data from Santé Publique France (SPF) (https://www.data.gouv.fr/fr/organizations/sante-publique-france). For all models, the time series were log-transformed, because the raw data is locally expected to demonstrate exponential growth behavior. The daily data was smoothed using a centered moving average over seven days.

**Table 1.** Surveillance data.

| Name | Source | Period | Fitting Windows | Time Resolution | Spatial Resolution (# regions) |
| --- | --- | --- | --- | --- | --- |
| COVID Cases, Hosp., Deaths DE | RKI | 2020-2023 | 140 | daily | Regional (16+1) |
| COVID Cases, Hosp., Deaths FR | SPF | 2020-2023 | 140 | daily | regional (13+1) |
| Influenza Cases DE | RKI | 2020-2024 | 30 | weekly | country (1) |
| SARI Hosp DE | RKI | 2014-2024 | 80 | weekly | country (1) |

### 2.2 Metadata

As metadata, we incorporated data from Google Trends following Wang et al. [24]. First, we identified the 20 top symptoms of COVID-19 which were used as search terms in Google Trends. By accessing their API (https://github.com/googleapis/google-api-python-client) we extracted the normalized daily number of counts each term was searched for. For smoothing we applied a centered moving average over seven days.

### 2.3 Base Models

In the following, we introduce the used base models and explain why they are suitable for time series forecasting. The exact training and tuning procedure is explained in section 2.5.

#### Linear Regression

Assuming that a pandemic follows exponential-like behavior - exponential growth and decay in waves, log-transforming the data will locally yield linear slopes. Therefore, linear regression can be used to fit linear models to the log-transformed data. Using the regression parameters the fit can then be extrapolated to estimate short-term forecasts. We used the scikit-learn library (version 1.0.2) “linear-model”.

#### ARIMA

Autoregressive integrated moving average (ARIMA) models use the statistical characteristics of stationary data. They are popular for time-series forecasting and have previously been applied to modeling of COVID-19 surveillance data [25–28]. A stationary series has no trends and consistently varies around its mean. That means short-term random time patterns can be extracted accordingly and used for forecasting. Here we employed a non-seasonal ARIMA model fitted to short-term periods which are not expected to show seasonal effects. This also applies to the Influenza and SARI data. Using seasonal ARIMA would only become effective when including at least two seasons. In this case, the ARIMA models depended on three parameters:

- *p* the number of autoregressive terms
- *d* the degree of differencing the data to make it stationary
- *q* the number of lagged forecast errors

With these parameters the general ARIMA forecasting equation is defined as:

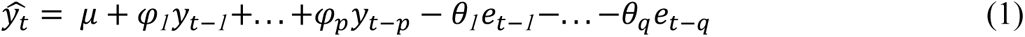

Here 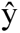 corresponds to the forecast which is computed as the deviation of the mean 𝜇 of a stationary time series with 𝜑, the slope parameters for each of the p previous values y, and q moving average parameters θ with autocorrelation errors e. This means the model learns to predict future steps based on the mean of a stationary time series with adjusted autocorrelation errors and a lagged period [20]. To ensure stationarity we employed the differencing technique. Differencing refers to the process of computing the differences between consecutive values in a time series. Doing so transforms the time series to the fluctuations of consecutive values, which in first or second order often leads to stationarity [29]. To find the best parameters (*p,d,q*) we implemented the auto-ARIMA functionality which is part of the pmdarima library (version 2.0.3) [30]. This essentially corresponds to a hyperparameter tuning.

#### Random Forest and XGBoost

Both Random Forest and eXtreme Gradient Boosting (XGBoost) are based on decision trees. However, they differ to a great extent in their training algorithm. Random Forest builds an unweighted ensemble of decision trees, which are - by applying bagging - trained in parallel on different subsets of the data and then averaged [22]. In contrast, XGBoost builds its decision trees one after the other and corrects the residual errors made by the previously trained weighted decision tree ensemble using gradient descent [21]. Both models are commonly applied to tabular data but have also been shown to be successful in time series forecasting [31,32], also for COVID-19 [33–36]. Since they are based on decision trees, they can only extrapolate based on previously seen training data. If the models are tasked to predict values outside of the training data, they will predict an average of this. Therefore, we first log-transformed the data and then applied the previously explained differencing technique [29] to ensure stationarity. We tested for stationarity by applying the augmented dickey-fuller (ADF) test [37]. Using stationary data does not only mean that the extrapolation problem is reduced, but also that it is possible to apply *k*-fold cross-validation for hyperparameter tuning since stationarity breaks the time dependence [38]. For Random Forest we used the scikit-learn library (version 1.0.2) “ensemble” and for XGBoost the xgboost library (version 1.7.3).

#### LSTM

Recurrent Neural Networks (RNNs) are commonly used for sequencing data. Their advantage compared to standard neural networks is their internal memory, i.e., their ability to remember and learn the influence of previous steps on current steps. Opposed to standard RNNs, a Long Short Term Memory (LSTM) can learn longer-range time patterns of time series without suffering from the vanishing gradient problem [23]. LSTMs have also been applied for time series forecasting in COVID-19 [39,40] Here we implemented an LSTM model in which the last hidden state - the state that contains the latent information about the time series - is decoded in a fully connected layer with output dimension according to the prediction window (14 days / 2 weeks). The LSTM model and fully connected layer were implemented using pytorch (version 1.11.0).

### 2.4 Sliding Window Approach for Model Training, Tuning, and Evaluation

We split the time series into N (140, 30, 80) training and testing windows. For this, we followed a sliding window approach (see Figure 1) with a training window size of 70 days for the data with daily resolution and 52 weeks for the data with weekly resolution, respectively. A testing window size of 14 days (daily data) / 2 weeks (weekly data), and a step size of 7 days (daily data) / 1 week (weekly data) were used. The objective was to forecast the value of the time series 14 days ahead of time, counted from the end of the training window. ARIMA, Random Forest, as well as XGBoost, were trained on the whole training set, and the log-linear regression was fitted on the last seven days (daily data) / five weeks (weekly data) of the training data. These models were applied separately for each region. For the LSTM model, however, we needed more data. Therefore, we applied another sliding window approach by creating fitting windows of size 7 days (daily data) / 5 weeks (weekly data) and evaluation windows of size 14 days (daily data) / 2 weeks (weekly data), with a stride of 1 day (daily data) / 1 week (weekly data). We decided to not train one LSTM model per region but to shuffle the regional windows, to increase the amount of training data. Therefore, the LSTM models’ training objective was to predict 14 days (daily data) / 2 weeks (weekly data) ahead based on 7 days (daily data) / 5 weeks (weekly data) of training data. We then tuned the hyperparameters of Random Forest, XGBoost, and LSTM models for each training window and region if applicable using Optuna (version 2.10.1). For more information about hyperparameter tuning, we refer to the supplementary materials (see S1). Since we were using the auto–ARIMA functionality, the hyperparameter tuning was done via a grid search, where the maximum parameter values for (*p,q*) were set to (14,14). Random Forest and XGBoost were tuned with an inner 5-fold cross-validation, while for the LSTM we split the training windows into 80% training and 20% validation sets. Here *k*-fold cross-validation was not possible, because we had to consider the time dependencies of the data. Also, we decided not to follow the classical time series split for time series cross-validation due to the increased run time and insufficient data. For more details we refer to [38]. After hyperparameter tuning we retrained the models - using the best hyperparameters for each fitting window - on the whole training data and progressively predicted and evaluated on the test windows.

**Fig 1:**
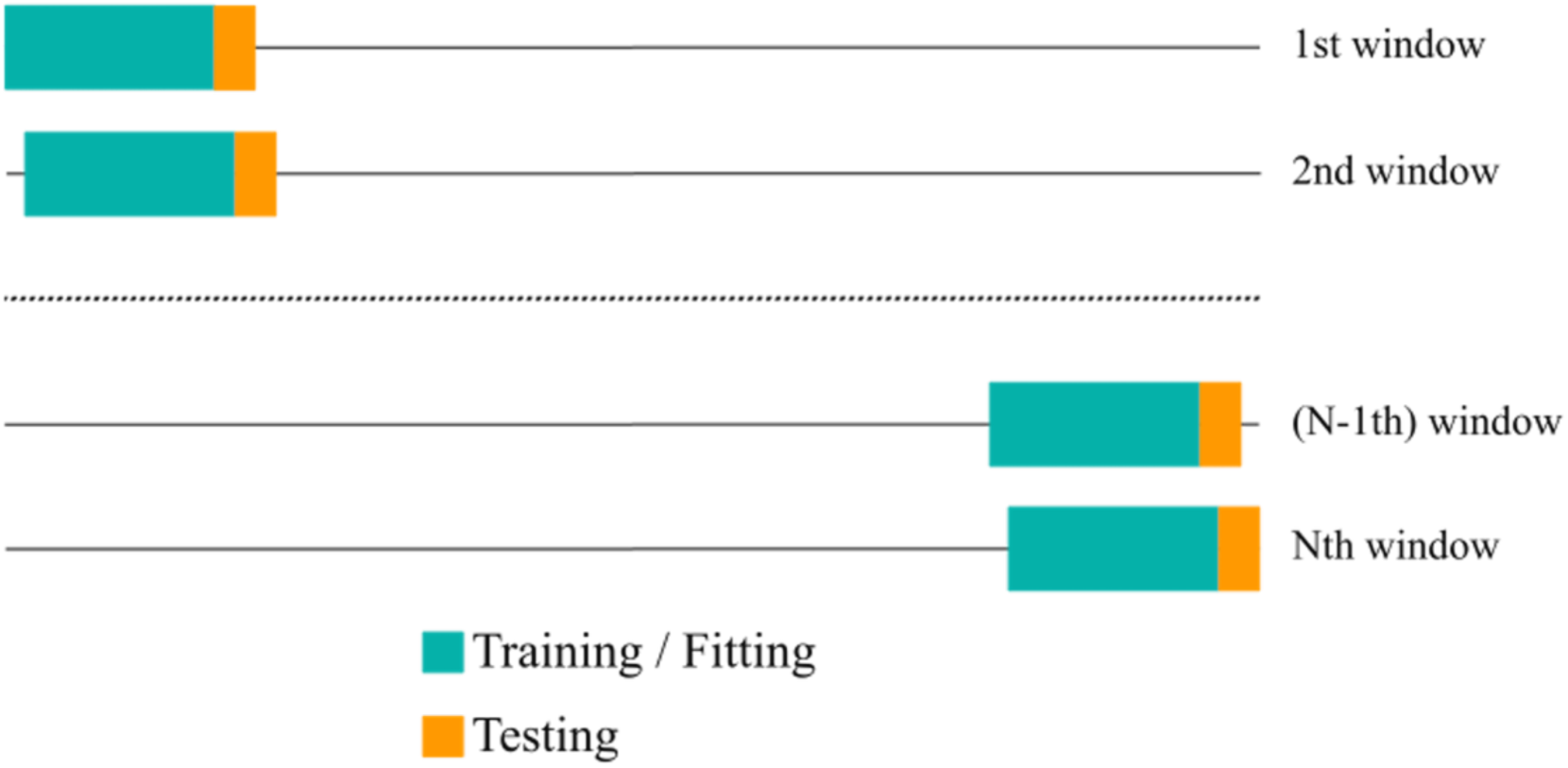
Sliding window approach. The time series was split into N training and testing windows. Models were tuned on the training windows using cross-validation (green), retrained with the best hyperparameters, and then forecasted 2 weeks ahead. Predictions were compared against real values observed in the test window (yellow).

### 2.5 Model Evaluation Metrics

To evaluate the performance of the base models and later the ensemble we used the mean absolute percentage error (MAPE) as a metric:

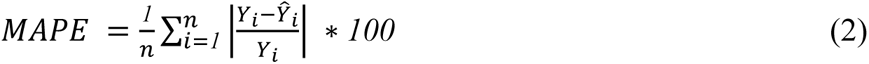

with *Y* as the real value, 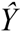 as the predicted value, and *n* as the number of data points, in our case the prediction window (14 days / 2 weeks). The MAPE represents the deviation of the prediction from the real data in percent and is therefore a more tangible measure than the mean squared error. The MAPE alone should not be used for determining the performance of a model, since it is scale-dependent [41]. However, it is a good measure to quantitatively compare the performances of different models.

### 2.6 Baseline Ensemble Approaches

As a baseline, we implemented two basic ensemble algorithms, more specifically the mean and the median of the model forecasts. Additionally, we built an ensemble algorithm that always chooses the model that performed best in the previous testing period (Prev.-Best). This corresponds to a first step in accounting for the dynamics of the pandemic and thus the dynamic performance of each base model.

### 2.7 Dynamic Model Selection and Stacking

We here propose two possible extensions of the baseline ensemble methods discussed before: i) dynamic model selection and ii.) dynamic model stacking, an extension of a classical stacked regressor approach [42]. In practice, we realize both approaches by training a meta-model, which we chose as a simple MLP architecture with a tunable hidden layer size. The input for the meta-model constituted of the predicted values as well as estimated prediction performances of all base models, by concatenating the MAPEs of the previous testing period to the log-transformed forecasts of the current testing period. Therefore, the MLP has five input vectors - one per base model. After each hidden layer a rectified linear unit (ReLU) activation function is applied. The output layer is designed to hold one node per model including a softmax head at the end. As mentioned above, there are two learning objectives:

1. Dynamic model selection: The meta-model is trained to always select the model with the highest softmax output. This essentially corresponds to a classification task, where the model with the highest probability score is selected.
2. Dynamic model stacking: The meta-model is trained to multiply the base model forecasts by the individual softmax inputs. These weighted outputs are then aggregated into one final ensemble output. This essentially corresponds to a weighted mean, since the softmax outputs add up to 1.

Both learning objectives are trained with a weighted MSE (WMSE) loss function:

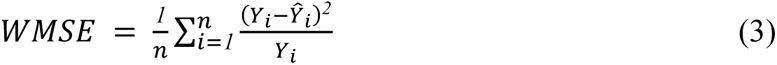

with *Y* as the real value, 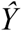 as the predicted value, and *n* as the number of data points, in our case the prediction window (14 days / 2 weeks). The WMSE has the advantage that it penalizes the relative deviation rather than the absolute deviation. For example: say the real value was 100 and the predicted value was 101 the MSE would be 1. It would be the same for the real value being 1 and the predicted value being 2. However, the relative deviation would be 1% vs. 100%. Weighting the MSE by the real values results in normalizing this error to the real value scale. In this case, the WMSE would be 0.01 and 1, respectively - the deviation of 100% is accordingly penalized much more than the deviation of 1%.

### 2.8 Inclusion of Metadata

Our above-described modeling approaches used only surveillance data, their forecasts, and estimated prediction performances as input for the base models and meta-model, respectively. In our previous publication, we showed that social media data is not only correlated with surveillance data but can also be used to forecast up- and downtrends of pandemic waves [24]. Therefore, we wanted to test if the inclusion of social media data or further metadata could improve the prediction performance of the meta-model. We employed Google Trends data as described above and applied a sliding window approach, where we used the past *n* (*n*=2,3,4) weeks (before the last date of our fitting period) as the training period for the metadata. To extract time patterns, we used an LSTM model and concatenated the last hidden state to the input of the meta-model, extending the input feature vector to include the forecasted value, the prediction performance estimated from the previous testing period, and now the information coming from the metadata. The meta-model was then trained in the same way as before, but now the weights of the LSTM were also updated according to the weighted MSE loss between the output and the real values. Due to the high computational burden and to be consistent with Wang et al. [24], we evaluated this approach on German surveillance data only.

### 2.9 Overall Ensemble Model Pipeline

The final ensemble model pipeline can be seen in Figure 2. The surveillance data, which was previously split into training and testing windows according to the sliding window approach explained above, is used as input for the base models. The tuned base models are trained in parallel and create a rolling forecast based on the testing windows. After each testing window, the baseline models (mean, median, Prev.-Best) are created. The predictions are evaluated using the current test data and the MAPE as a metric. The base models’ forecasts together with their performance on the previous testing period are concatenated to form the input vectors of the meta-model. If metadata is included the metadata is fed into an LSTM model, of which the last hidden state - the latent representation of the metadata - is concatenated to the input vectors of the meta-model. As described in Section 2.4 the overall data was split into 80% training and 20% test. The training data was further split into 5 folds for an inner 5-fold cross-validation and hyperparameter tuning (see S1). Finally, the models’ performances over the test data were averaged and an output containing these mean performances was returned. The code for the ensemble model can be accessed on Git Hub (https://github.com/SCAI-BIO/Dynamic_Ensemble).

**Fig 2:**
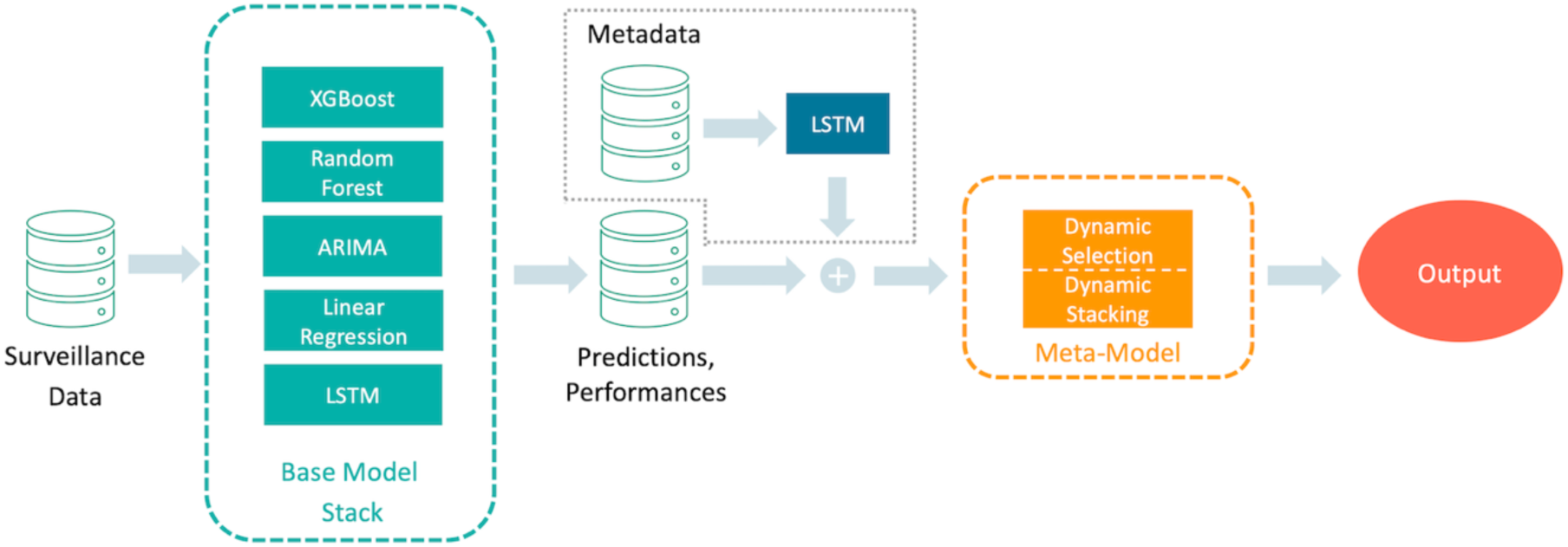
Overall ensemble Model Pipeline. The surveillance data is fed into the base models which produce forecasts. All forecasts and their evaluation (plus the latent representation of the metadata) are used as input for the meta-model which outputs forecasts either based on dynamic selection or dynamic stacking.

### 2.10 Model Ranking and Post-Hoc Analysis

To quantitatively compare the model performances across datasets we first ranked all models according to a consensus ranking [43] - allowing for ties - based on Kemeny’s axiomatic approach [44]. The algorithm compares models pairwise and counts how often one model is ranked above the other. The total sum of counts is then used to form the consensus ranking. This ranking alone, however, does not necessarily mean that one model’s performance is significantly different from another model. To test for statistical significance across models we thus used a Kruskal-Wallis test [45]. To test which individual models differed significantly from each other we then used a pairwise Wilcoxon test as post-hoc test [46]. All p-values are adjusted for multiple testing based on the Holm-Bonferroni method [47]. Statistical tests were implemented using R (version 4.3.0) and the libraries ConsRank (version 2.1.4) and stats (version 4.3.0).

## 3. Results

In the following, we display the results on the country level and the regional results aggregated (mean over all regional results) at the country level. The complete set of results can be found in the supplementary material. Model performances are displayed as the mean MAPE of all test windows in percent together with its standard error in parentheses. We use the following abbreviations for the models: Linear Regression - LR, XGBoost- XG, Random Forest - RF, and ensemble baseline by best model selection - Prev.-Best. Since the Influenza dataset only contains 30 test windows (and just 6 test windows for the meta-model) the results should be interpreted cautiously, as the small sample size leads to a reduced statistical meaningfulness. Additionally, we provide the results from the consensus ranking and the Kruskal-Wallis as well as the Wilcoxon test. For all results, the Kruskal-Wallis test returned a significant p-value of less than 5%.

### 3.1 Base Models versus Baseline Ensembles

First, we evaluated the base and baseline ensemble models by computing and testing a rolling forecast over the full time series. This resulted in 140 test windows for the daily COVID-19 datasets, 30 test windows for the weekly Influenza cases, and 80 test windows for the weekly SARI hospitalization. The results are summarized in Table 2. For a better overview, we colored the three best models for each dataset / dataset aggregation. First, looking at the base models’ performances on the daily COVID-19 datasets, it can be seen that mostly linear regression and ARIMA performed best and LSTM and XGBoost worst. On the weekly dataset, Random Forest and XGBoost were able to perform similarly well as ARIMA. Here the linear regression showed a reduced performance. Taking a look at the baseline ensemble methods shows that mean and median baseline ensembles rarely performed as one of the three best models, but the Prev.- Best method was able to outperform most of the base models in many instances; at least for the daily COVID-19 datasets. Evaluated on the SARI hospitalization dataset, mean managed to be the best model. Since Prev.-Best always selects the best model of the previous week, we integrated a counter to keep track of this selection. The number of model selections per dataset can be seen in Figure 3. This agrees with the results displayed in Table 2. The base models that performed best were also the ones being selected most often. But, still, the other base models were selected a considerable amount of times. Finally, it can be observed that the variance of the Prev.-Best method performances tended to be smaller than the variance of the base models (perhaps excluding ARIMA) performances. Now looking at the consensus ranking, we can see that ARIMA and Prev.-Best were both ranked first, followed by the mean and median baseline ensemble methods. Table 3 shows the p-values computed by the pairwise Wilcoxon test. It can be seen that indeed no significant difference between the Prev.-Best method and ARIMA could be found. However, they were both found to be significantly different than all other methods.

**Fig 3:**
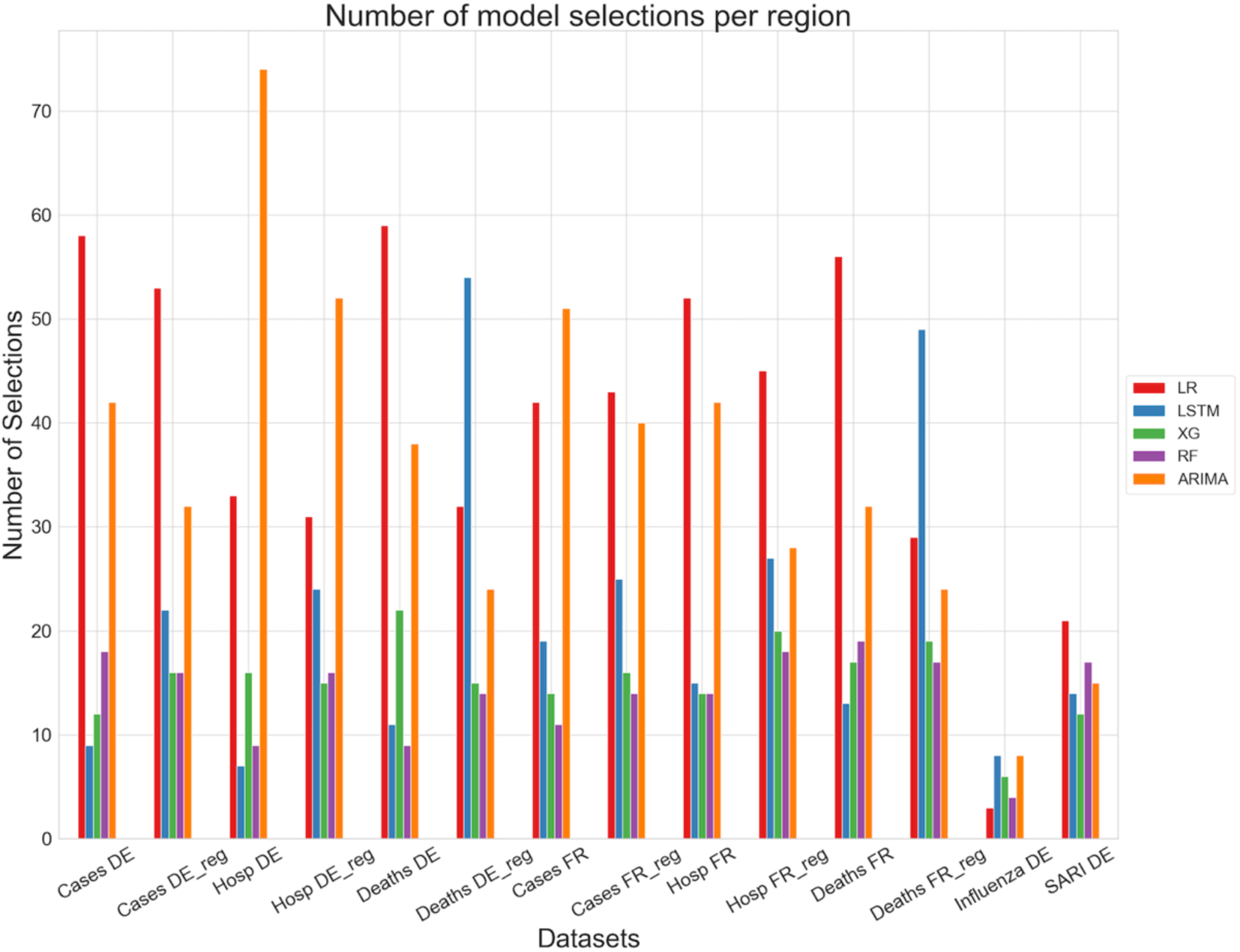
Number of model selections per region bar plot. DE (FR) stands for German (France) country level and DE_reg (FR_reg) for German (France) regional level aggregated to country level.

**Table 2:**
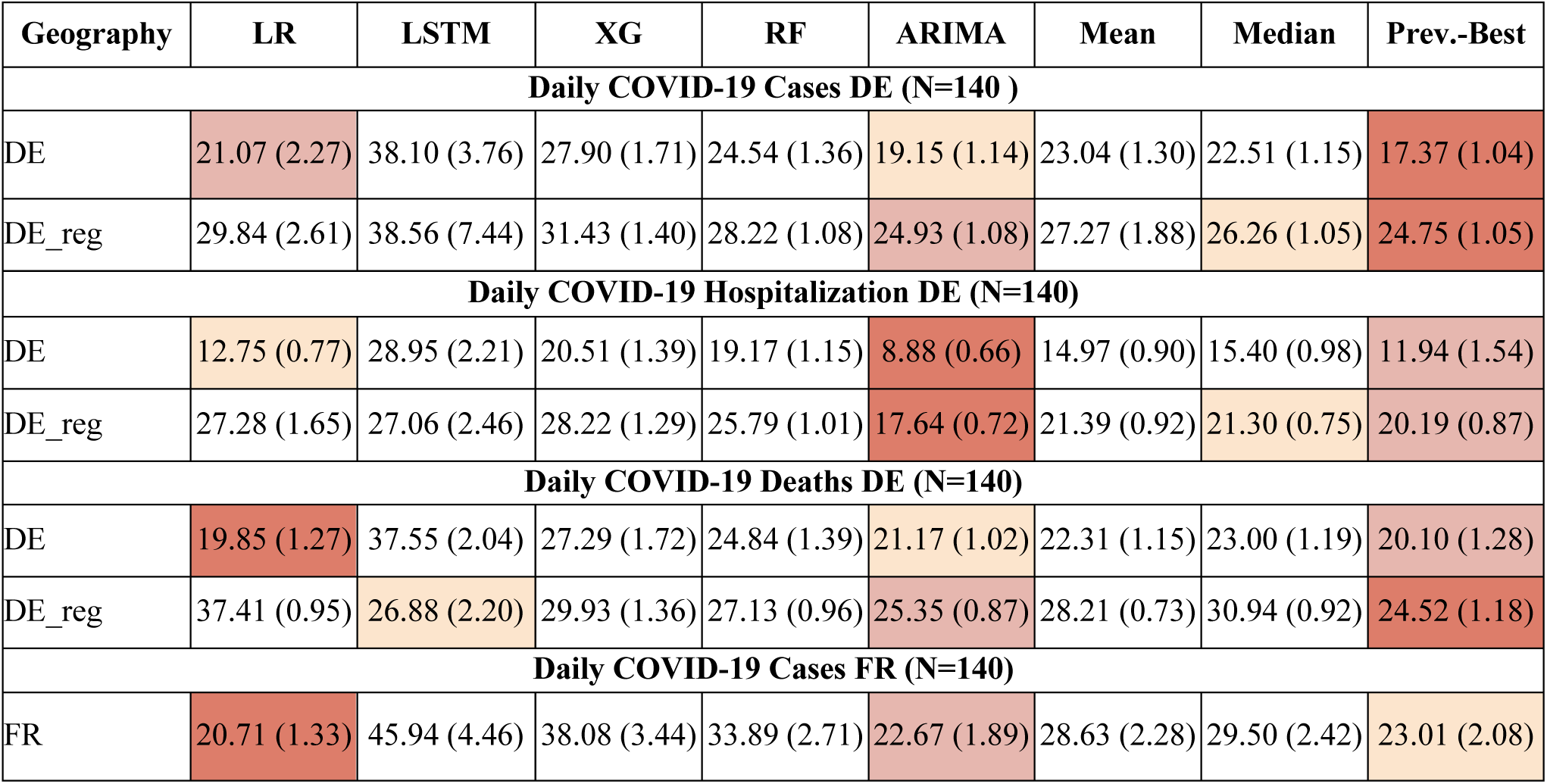
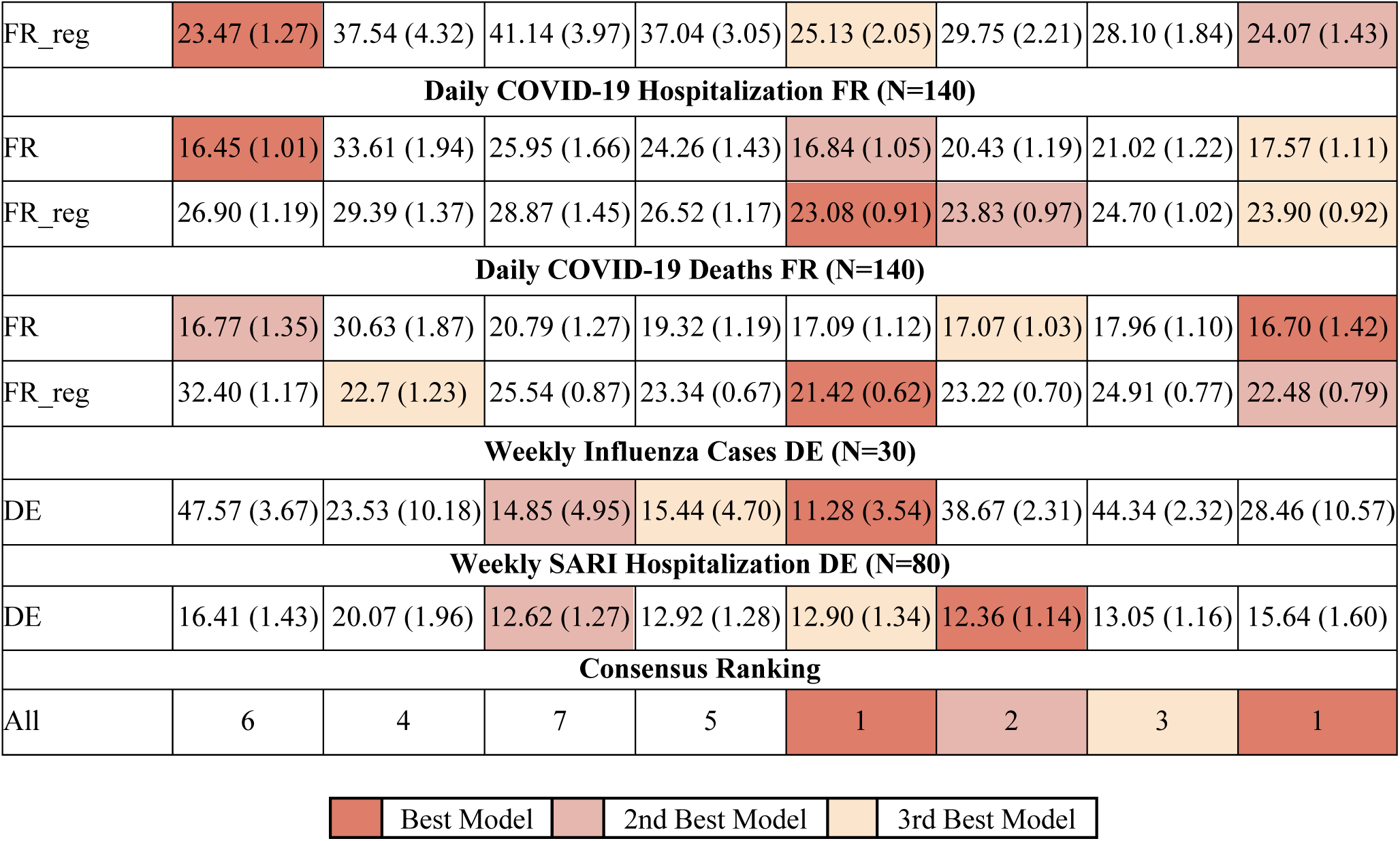
Base models versus baseline ensemble methods. The performances are given as the mean MAPE and its standard error in parentheses of the N test windows for each dataset / dataset aggregation. The best three models are colored according to the provided legend. DE (FR) stands for German (France) country level and DE_reg (FR_reg) for German (France) regional level aggregated to country level. The last two rows display the results from the consensus ranking over all datasets and models.

**Table 3:** Base models versus baseline ensemble approaches: pairwise Wilcoxon Test (adjusted p-values).

|  | ARIMA | LR | LSTM | Mean | Median | Prev. Best | RF |
| --- | --- | --- | --- | --- | --- | --- | --- |
| **LR** | <1E-16 | - | - | - | - | - | - |
| **LSTM** | <1E-16 | 6.59E-02 | - | - | - | - | - |
| **Mean** | <1E-16 | 7.55E-14 | 4.58E-06 | - | - | - | - |
| **Median** | <1E-16 | 8.46E-03 | 8.89E-01 | 7.32E-06 | - | - | - |
| **Prev. Best** | 8.42E-01 | <1E-16 | <1E-16 | <1E-16 | <1E-16 | - | - |
| **RF** | <1E-16 | 8.89E-01 | 2.01E-01 | 1.03E-11 | 9.41E-02 | <1E-16 | - |
| **XG** | <1E-16 | 5.90E-04 | 1.07E-09 | <1E-16 | 1.03E-11 | <1E-16 | 2.14E-05 |

**Table 4:**
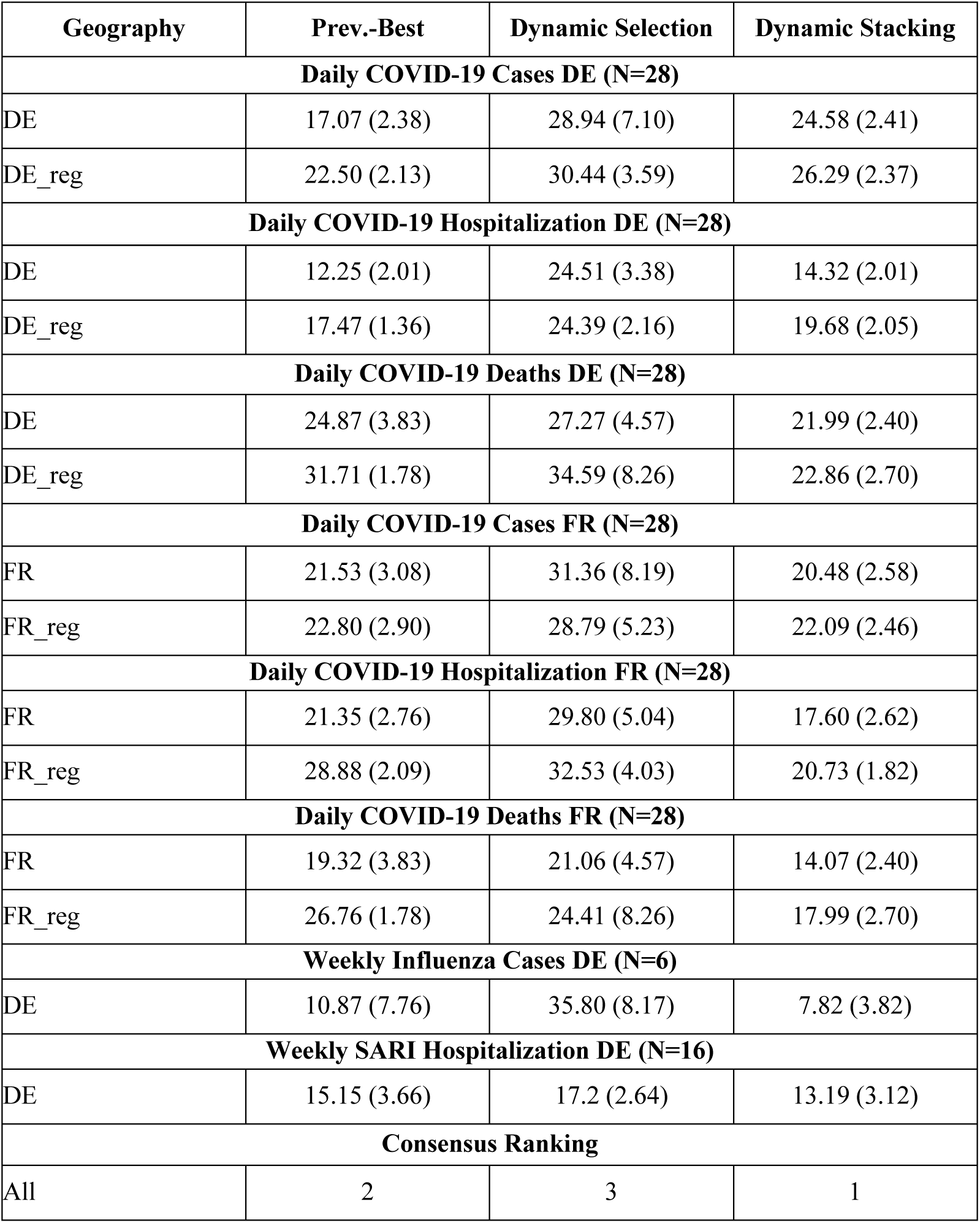
Comparison of ensemble modeling approaches. The performances are given as the mean MAPE and its standard error in parentheses of the N test windows for each dataset / dataset aggregation. DE (FR) stands for German (France) country level and DE_reg (FR_reg) for German (France) regional level aggregated to country level.

### 3.2 Baseline Ensembles versus Dynamic Model Stacking and Selection

Next, we evaluated our proposed Dynamic Model Stacking and Selection approaches against the previously tested Prev.-Best method. Since the meta-model was trained on 80% of the test windows, the number of test windows for the meta-model was reduced to 28 for the daily COVID-19 dataset, 6 for the weekly Influenza cases, and 16 for the weekly SARI hospitalization. According to the results presented in Table 3 Dynamic Selection was not able to outperform Prev.-Best. However, Dynamic Model Stacking outperformed Prev-Best and Dynamic Model Selection on the French and German COVID-19 deaths datasets and was the second-best model on the German COVID-19 hospitalization dataset. Moreover, it outperformed Dynamic Model Selection and Prev.-Best on the weekly datasets. Also, the variance of the Dynamic Model Stacking approach tended to be reduced compared to the other methods. The results of the consensus ranking are again in line with the findings above. The Dynamic Model Stacking method is ranked first being significantly better than Prev.-Best (see Table 5).

**Table 5:** Prev.-Best versus Dynamic Model Stacking and Selection: pairwise Wilcoxon Test (adjusted p-values).

|  | Prev.-Best | Dynamic Selection | Dynamic Stacking |
| --- | --- | --- | --- |
| Dynamic Selection | 3.24E-01 | - | - |
| Dynamic Stacking | 2.57E-07 | 2.17E-10 | - |

For the sake of completeness, we also list in Table S3.2.1 an additional comparison of Dynamic Model Stacking against all base models, which have been re-trained on the same dataset. Also, in this comparison, Dynamic Model Stacking could significantly outperform ARIMA as the best base model (p = 5.74E-3).

### 3.3 Potential Benefits of Including Metadata

Finally, we wanted to assess whether the inclusion of metadata - here Google Trends symptom counts - could further enhance our proposed Dynamic Model Stacking approach. We could not find an improvement by including metadata. Both Dynamic Model Stacking with and without metadata were ranked on position after consensus ranking and the Wilcoxon-test showed no significant differences between both approaches.

## 4. Discussion

The COVID-19 pandemic highlighted the need for robust models that can accurately forecast the spread of the pandemic and can adjust dynamically to external factors such as newly imposed non-pharmaceutical interventions, new virus variants, vaccination, seasonal effects, and others. In this work, we initially tested and compared different base machine learning models against basic ensemble methods (mean, median, Prev.-Best), demonstrating no statistically significant benefit of these simple techniques compared to a state-of-the art ARIMA time series forecasting model. Only Prev.-Best was found to perform en par with ARIMA. Even though the other base models did not perform as well as ARIMA we still found them to be frequently selected as significantly often as being the best model in the previous test window. Therefore, we decided to keep these other base models in the model ensemble.

We then developed a meta-model that can either dynamically select one of the base model predictions or add the weighted base model predictions into one forecast. Interestingly, only Dynamic Model Stacking turned out to outperform Prev.-Best while at the same time showing a reduced variance in prediction performance. The inclusion of Google Trends symptom counts as metadata could not further improve Dynamic Model Stacking significantly and thus cannot be recommended.

Comparing the performances on the country and regional level we found Dynamic Model Stacking on the country level to be superior. We assume that this comes from the data quality of the regional data. Since we are working with surveillance data that needs to be registered at local health departments, it can happen that mistakes are made on a regional level. These mistakes would have reduced effects when the regional data is aggregated to the country-level data.

In general, we saw considerable differences between model performances on the daily and the weekly datasets. Specifically, the SARI hospitalization data suggests that decision tree-based models - especially Random Forest - and the mean and median baseline ensemble methods work well here. In this regard, we should point out that in the weekly data, the task is just to forecast the next two data points (2 weeks) which seems to be handled well by decision tree models. Linear regression struggles here because the model is fit to the past 5 data points (i.e. weeks), hence resulting in over-smoothing.

We should mention the limitations of the non-COVID datasets, specifically limited sample size and, in case of the Influenza, also seasonal fluctuations (see Figure S2). Moreover, the SARI dataset contains hospitalization due to different pathogens. These limitations lead to non-trivial challenges for learning a good model.

A comparison of our findings with those in other studies is challenging because different datasets (perhaps even just one wave rather than a whole pandemic), different forecasting horizons, and different metrics have been used. Paireau et al. [11] developed an ensemble model (mean) and forecasted among other indicators the COVID-19 hospitalization in France. On country-level data, they documented a mean MAPE of 20% and on aggregated regional level of 30% for a 14-day forecast horizon. Our best model - evaluated on the French COVID-19 hospitalization dataset - achieved a mean MAPE of around 17% and around 20% to 23% (for the meta-model test windows or all test windows) on country-level and regional aggregated country-level data, respectively. Heredia Cacha et al. [10] forecasted COVID-19 cases in Spain using different ensemble methods (mean, median, weighted average) and documented a mean MAPE of around 30% for a 14-day forecasting horizon. We achieved a MAPE of around 17% to 25% for forecasting the number of COVID-19 cases in Germany and France. Stating that our models are better than the ones of Paireau et al. and Heredia Cacha et al. would not be fair, though, since we are not using the exact same data. However, this comparison confirms that our models are generally competitive with others reported in the literature.

## 5. Conclusion

A major challenge for the modeling of pandemic situations, specifically COVID-19, is their highly dynamic character. Rapid introduction of non-pharmaceutical interventions, newly emerging virus variants, vaccinations, and seasonal effects strongly violate the typical assumption of stationarity in time series modeling and forecasting and thus negatively affect the generalization ability of models. In this regard, we here proposed a novel ensemble learning strategy, in which a meta-model learns to dynamically weigh and integrate a set of base models based on currently observed data and past performance indicators. Based on results from 8 datasets, our Dynamic Model Stacking approach was able to outperform state-of-the art time series forecasting techniques, such as ARIMA, and other ensemble learning approaches. Furthermore, we could show that our method could not be further improved by adding further metadata, such as Google searches.

Of course, our proposed Dynamic Model Stacking approach is not without limitations. Most importantly, machine learning methods need a sufficient amount of training data, i.e. retrospective pandemic data, which are not always available at the beginning of a pandemic. A potential strategy in future pandemics might thus be to start building a collection of comparable simple base models, specifically including ARIMA, and then to train Dynamic Model Stacking once sufficient historical data is available.

While we only evaluated Dynamic Model Stacking on surveillance data of COVID-19, SARI, and Influenza, our method is not limited per se to these data. Dynamic Model Stacking could potentially be applied also to other areas, where non-stationary time series forecasting plays a role, e.g. traffic, energy consumption, air flights, and others. Moreover, future work could apply Dynamic Model Stacking to age-distributed data, especially to the vulnerable, mostly elderly population.

## Data Availability

The data is publicly available under the following three links: https://github.com/robert-koch-institut https://github.com/googleapis/google-api-python-client https://www.data.gouv.fr/fr/organizations/sante-publique-france/#/presentation

https://github.com/robert-koch-institut

https://www.data.gouv.fr/fr/organizations/sante-publique-france/#/presentation

## Acknowledgments

This work has been supported by the AIOLOS (Artificial Intelligence Tools for Outbreak Detection and Response) project. The project was supported by the French State and the German Federal Ministry for Economic Affairs and Climate Action (grant number 01MJ22005A) and the French Ministry of Economy and Finance in the context of the France 2030 initiative and the Franco-German call on Artificial Intelligence technologies for risk prevention, crisis management, and resilience.

## Author Contribution

Conceptualization, methodology, supervision, project administration, and funding acquisition: HF. Data curation, formal analysis, visualization, investigation, validation, and writing—original draft: JB, DV, JG, HF Writing—review and editing: JB, DV, JG, and HF. All authors contributed to the article and approved the submitted version.

## Supporting Informations

**S1 Table. Hyperparameters for base models and meta-models.**

(PDF)

**S2 Figure. Comparison SARI and Influenza.** SARI and Influenza incidence from May 2022 to May 2023.

(PNG)

**S3 Appendix. Complete results**

(PDF)

