## Supplementary material for "A dynamic ensemble model for short-term forecasting in pandemic situations": S1_Table.pdf

| Model | Hyperparameter | Search Space | Sampler |
|---|---|---|---|
| Random Forest | Min_samples_split<br>Min_samples_leaf | [2,4,8,16]<br>[1,2,4] | Grid-Search |
| XGBoost | Learning rate<br>Max_depth<br>Reg_lambda<br>Reg_alpha | (0.01,0.5)<br>(2,12)<br>(0,10)<br>(0,10) | TPE |
| LSTM | Learning rate<br>Batch size<br>Hidden state dim.<br># Hidden layers<br>Droprate | [0.001,0.005,0.01,0.05,0.1]<br>[16,32,64,128]<br>[16,32,64,128]<br>[1,2,3]<br>[0.0,0.2,0.5] | TPE |
| Meta-Model | Learning rate<br>Batch size<br># Hidden layers<br>Hidden state dim. | [0.001,0.005,0.01,0.05,0.1]<br>[16,32,64,128]<br>[0,1,2]<br>[8,16,32] | Grid Search |
| Meta-Model+LSTM* | Lookback Window<br>Learning rate<br>Batch size<br># Hidden layers<br>Hidden state dim.<br>Learning rate*<br>Hidden state dim.*<br># Hidden layers*<br>Droprate* | [2,3,4]<br>[0.001,0.005,0.01,0.05,0.1]<br>[16,32,64,128]<br>[0,1,2]<br>[8,16,32]<br>[0.001,0.005,0.01,0.05,0.1]<br>[4,8,16,32]<br>[1,2,3]<br>[0.0,0.2,0.5] | TPE |

**Table S1: Hyperparameters for base models and meta-models.** The brackets correspond to discrete values, while the parentheses correspond to continuous parameter ranges. Hyperparameters denoted by a star belong to the LSTM model. The lookback window refers to the metadata. Hyperparameters were either optimized by using a grid-search or with a tree-structured Parzen estimator from optuna. As the base model's hyperparameters were tuned for each fitting window and dataset and the meta-model's hyperparameter for each dataset, we did not provide the final choices.
