## Supplementary material for "A dynamic ensemble model for short-term forecasting in pandemic situations": S3_Appendix.pdf

In the following we are displaying the regional results and aggregated results that were not included in the manuscript.

#### S3.1 Base Models versus Baseline Ensembles Results

| Geography | LR | LSTM | XG | RF | ARIMA | Mean | Median | Prev.-Best |
|---|---|---|---|---|---|---|---|---|
| DE | 21.07 (2.27) | 38.1 (3.76) | 27.9 (1.71) | 24.54 (1.36) | 19.15 (1.14) | 23.04 (1.3) | 22.51 (1.15) | 17.37 (1.04) |
| DE.BB | 29.02 (2.55) | 40.62 (9.78) | 32.43 (1.94) | 28.89 (1.64) | 27.87 (1.84) | 28.39 (2.55) | 27.14 (1.55) | 26.02 (1.65) |
| DE.BE | 27.34 (2.75) | 32.5 (4.56) | 28.93 (1.62) | 27.29 (1.36) | 24.58 (1.47) | 25.0 (1.66) | 25.07 (1.34) | 23.58 (1.68) |
| DE.BW | 24.17 (1.57) | 29.32 (1.73) | 31.57 (1.99) | 28.3 (1.71) | 24.18 (1.43) | 24.5 (1.39) | 26.38 (1.46) | 23.51 (1.57) |
| DE.BY | 22.71 (1.53) | 29.52 (1.61) | 30.08 (1.81) | 27.75 (1.53) | 22.8 (1.33) | 23.86 (1.25) | 25.12 (1.34) | 21.33 (1.35) |
| DE.HB | 32.03 (2.69) | 60.25 (19.64) | 32.85 (2.4) | 28.98 (1.84) | 25.98 (1.52) | 32.03 (4.06) | 27.07 (1.44) | 27.29 (1.78) |
| DE.HE | 24.37 (2.44) | 30.6 (2.18) | 29.91 (1.74) | 27.65 (1.49) | 23.07 (1.24) | 24.24 (1.39) | 25.86 (1.4) | 21.47 (1.65) |
| DE.HH | 27.6 (3.23) | 36.18 (7.15) | 27.74 (1.97) | 25.62 (1.64) | 21.37 (1.42) | 24.87 (1.95) | 23.28 (1.41) | 27.67 (6.0) |
| DE.MV | 35.76 (4.84) | 46.2 (12.82) | 30.97 (2.1) | 27.68 (1.89) | 25.35 (1.52) | 29.07 (2.99) | 26.71 (1.74) | 28.76 (4.43) |
| DE.NI | 24.79 (2.43) | 27.81 (1.7) | 27.04 (1.62) | 25.22 (1.4) | 22.05 (1.36) | 22.11 (1.27) | 23.47 (1.38) | 21.56 (1.34) |
| DE.NW | 29.74 (8.29) | 28.13 (1.76) | 29.73 (1.89) | 26.58 (1.52) | 21.26 (1.1) | 24.35 (2.19) | 24.97 (1.44) | 20.91 (1.33) |
| DE.RP | 27.15 (1.92) | 34.38 (4.22) | 32.19 (2.08) | 29.51 (1.77) | 25.05 (1.49) | 26.09 (1.72) | 27.01 (1.63) | 24.79 (1.6) |
| DE.SH | 31.57 (3.08) | 32.86 (4.53) | 29.78 (1.66) | 27.17 (1.35) | 23.6 (1.64) | 25.5 (1.65) | 25.35 (1.38) | 25.08 (2.58) |
| DE.SL | 35.54 (4.21) | 61.55 (21.01) | 37.35 (2.78) | 32.94 (2.16) | 27.93 (1.85) | 35.6 (4.68) | 30.77 (1.93) | 30.06 (2.33) |
| DE.SN | 47.42 (18.74) | 36.84 (6.56) | 36.37 (2.78) | 31.0 (2.14) | 28.77 (1.84) | 32.68 (4.64) | 28.32 (1.68) | 22.86 (1.47) |
| DE.ST | 32.76 (3.73) | 42.95 (11.68) | 34.22 (3.06) | 29.04 (2.28) | 28.63 (2.53) | 29.7 (3.19) | 27.87 (2.04) | 28.38 (2.87) |
| DE.TH | 25.55 (1.72) | 47.29 (17.87) | 31.79 (2.21) | 27.85 (1.64) | 26.45 (1.71) | 28.3 (3.93) | 25.83 (1.57) | 22.72 (1.52) |

**Table S3.1.1:** Base models versus baseline ensemble methods for COVID-19 cases in Germany. The performances are given as the mean MAPE and its standard error in parentheses. The regions are given in HASC Codes (http://www.statoids.com/ihasc.html)

| Geography | LR | LSTM | XG | RF | ARIMA | Mean | Median | Prev.-Best |
|---|---|---|---|---|---|---|---|---|
| DE | 12.75 (0.77) | 28.95 (2.12) | 20.51 (1.39) | 19.17 (1.15) | 8.88 (0.66) | 14.97 (0.9) | 15.4 (0.98) | 11.94 (1.54) |
| DE.BB | 33.15 (4.18) | 26.85 (2.45) | 29.31 (2.14) | 26.72 (2.03) | 19.86 (1.31) | 23.69 (1.81) | 23.05 (1.62) | 19.81 (1.5) |
| DE.BE | 21.96 (1.63) | 19.63 (2.04) | 20.98 (1.23) | 19.68 (1.13) | 14.16 (0.93) | 16.44 (1.01) | 16.34 (1.0) | 16.09 (1.1) |
| DE.BW | 17.54 (1.24) | 22.24 (1.25) | 26.73 (2.54) | 22.41 (1.42) | 13.58 (0.94) | 18.13 (1.11) | 18.99 (1.08) | 15.56 (0.89) |
| DE.BY | 16.41 (1.08) | 22.14 (1.36) | 24.97 (1.81) | 22.07 (1.4) | 11.47 (0.61) | 16.89 (0.99) | 17.8 (1.01) | 12.55 (0.68) |
| DE.HB | 49.6 (12.66) | 60.83 (21.57) | 37.83 (2.99) | 34.91 (2.58) | 25.5 (1.99) | 35.48 (5.18) | 28.74 (1.82) | 34.81 (5.25) |
| DE.HE | 16.8 (1.11) | 20.84 (1.26) | 23.24 (1.56) | 21.89 (1.42) | 11.97 (0.69) | 16.5 (0.97) | 17.37 (0.98) | 13.7 (0.85) |
| DE.HH | 31.77 (3.17) | 27.91 (2.55) | 28.35 (2.33) | 26.4 (2.19) | 20.01 (1.5) | 22.91 (1.68) | 23.11 (1.66) | 19.99 (1.17) |
| DE.MV | 34.03 (3.51) | 27.71 (2.5) | 31.76 (3.06) | 30.92 (3.23) | 24.45 (2.33) | 24.38 (1.73) | 25.65 (1.84) | 23.49 (1.87) |
| DE.NI | 22.03 (1.54) | 21.0 (1.24) | 23.35 (1.4) | 21.78 (1.32) | 14.99 (1.14) | 17.21 (1.03) | 18.15 (1.08) | 16.14 (1.08) |
| DE.NW | 14.9 (0.92) | 23.85 (1.48) | 21.96 (1.55) | 20.08 (1.31) | 10.48 (0.64) | 15.8 (0.95) | 16.85 (1.08) | 12.22 (0.75) |
| DE.RP | 26.81 (3.59) | 22.09 (1.24) | 26.54 (1.77) | 23.94 (1.54) | 17.37 (1.25) | 19.44 (1.29) | 19.82 (1.27) | 18.81 (1.38) |
| DE.SH | 28.87 (3.1) | 27.57 (4.39) | 28.66 (3.12) | 27.63 (3.11) | 18.99 (1.97) | 21.7 (2.13) | 21.92 (1.86) | 22.96 (2.66) |
| DE.SL | 40.95 (4.66) | 38.26 (5.36) | 37.65 (2.54) | 34.16 (2.13) | 23.2 (1.25) | 29.39 (1.89) | 28.08 (1.53) | 34.72 (5.07) |
| DE.SN | 26.3 (3.1) | 25.44 (1.83) | 33.55 (4.77) | 28.2 (2.62) | 17.63 (1.16) | 21.95 (1.46) | 22.16 (1.3) | 21.13 (1.43) |
| DE.ST | 28.89 (2.96) | 25.2 (2.2) | 30.4 (2.17) | 27.61 (1.78) | 18.81 (1.3) | 22.35 (1.4) | 22.35 (1.46) | 21.4 (1.65) |
| DE.TH | 26.5 (2.78) | 21.36 (1.38) | 26.2 (1.61) | 24.26 (1.36) | 19.82 (1.45) | 20.04 (1.17) | 20.39 (1.19) | 19.62 (1.18) |

**Table S3.1.2:** Base models versus baseline ensemble methods for COVID-19 hospitalization in Germany. The performances are given as the mean MAPE and its standard error in parentheses. The regions are given in HASC Codes (http://www.statoids.com/ihasc.html).

| Geography | LR | LSTM | XG | RF | ARIMA | Mean | Median | Prev.-Best |
|---|---|---|---|---|---|---|---|---|
| DE | 19.85 (1.27) | 37.55 (2.04) | 27.29 (1.72) | 24.84 (1.39) | 21.17 (1.02) | 22.31 (1.15) | 23.0 (1.19) | 20.1 (1.28) |
| DE.BB | 35.74 (1.64) | 24.02 (3.15) | 26.41 (1.92) | 24.54 (1.52) | 23.66 (1.57) | 27.64 (1.31) | 30.48 (1.52) | 23.32 (1.63) |
| DE.BE | 40.78 (2.84) | 26.75 (3.18) | 30.11 (2.36) | 26.27 (1.89) | 25.06 (1.63) | 27.68 (1.64) | 30.02 (1.53) | 26.28 (3.07) |
| DE.BW | 28.63 (1.62) | 28.26 (1.81) | 33.38 (2.34) | 30.11 (1.91) | 28.13 (1.64) | 27.48 (1.4) | 30.01 (1.54) | 25.17 (1.63) |
| DE.BY | 30.99 (1.84) | 29.2 (1.95) | 33.51 (2.16) | 30.19 (1.81) | 27.63 (1.84) | 27.33 (1.47) | 29.16 (1.52) | 24.24 (1.6) |
| DE.HB | 46.63 (2.01) | 28.59 (5.19) | 25.66 (1.9) | 22.97 (1.39) | 20.4 (1.22) | 29.82 (1.15) | 35.56 (1.17) | 26.87 (4.63) |
| DE.HE | 30.87 (2.33) | 27.24 (1.82) | 32.66 (2.16) | 28.69 (1.73) | 26.53 (1.58) | 25.85 (1.3) | 27.99 (1.46) | 26.04 (2.22) |
| DE.HH | 41.65 (2.33) | 25.18 (2.41) | 28.91 (1.81) | 26.94 (1.5) | 24.38 (1.29) | 28.4 (1.17) | 30.65 (1.27) | 26.27 (2.17) |
| DE.MV | 43.37 (2.42) | 23.65 (3.64) | 24.75 (2.02) | 23.73 (1.73) | 21.9 (1.66) | 29.67 (1.31) | 32.51 (1.43) | 20.42 (1.88) |
| DE.NI | 36.79 (4.08) | 25.04 (1.52) | 31.14 (2.13) | 29.27 (1.9) | 25.79 (1.55) | 26.6 (1.51) | 28.37 (1.46) | 23.78 (1.43) |
| DE.NW | 27.17 (1.82) | 30.11 (1.95) | 29.02 (1.76) | 26.88 (1.4) | 23.81 (1.3) | 24.03 (1.22) | 26.34 (1.3) | 23.89 (1.39) |
| DE.RP | 37.39 (1.95) | 28.08 (2.87) | 33.77 (2.98) | 30.08 (2.0) | 27.61 (1.9) | 29.65 (1.54) | 32.08 (1.52) | 25.21 (1.72) |
| DE.SH | 40.16 (2.41) | 25.64 (3.5) | 27.71 (2.11) | 24.92 (1.52) | 22.56 (1.4) | 27.62 (1.3) | 29.71 (1.38) | 22.11 (1.5) |
| DE.SL | 43.31 (2.36) | 29.46 (4.43) | 27.08 (1.94) | 25.34 (1.63) | 23.13 (1.55) | 29.71 (1.35) | 32.97 (1.36) | 25.14 (1.99) |
| DE.SN | 37.25 (2.14) | 28.69 (2.68) | 33.36 (2.6) | 29.02 (1.9) | 32.1 (3.88) | 30.92 (1.52) | 33.24 (1.61) | 25.23 (1.81) |
| DE.ST | 41.44 (2.62) | 24.22 (2.71) | 28.74 (2.05) | 26.11 (1.63) | 25.81 (1.58) | 29.69 (1.34) | 33.07 (1.43) | 24.9 (2.89) |
| DE.TH | 36.35 (2.07) | 25.93 (2.62) | 32.59 (2.3) | 29.07 (1.79) | 27.14 (1.83) | 29.28 (1.29) | 32.86 (1.43) | 23.49 (2.13) |

**Table S3.1.3:** Base models versus baseline ensemble methods for COVID-19 deaths in Germany. The performances are given as the mean MAPE and its standard error in parentheses. The regions are given in HASC Codes (http://www.statoids.com/ihasc.html).

| Geography | LR | LSTM | XG | RF | ARIMA | Mean | Median | Prev.-Best |
|---|---|---|---|---|---|---|---|---|
| FR | 20.71 (1.33) | 45.94 (4.46) | 38.08 (3.44) | 33.89 (2.71) | 22.67 (1.89) | 28.63 (2.28) | 29.50 (2.42) | 23.01 (2.08) |
| FR.AC | 24.38 (1.88) | 31.43 (2.18) | 40.41 (4.81) | 37.92 (4.04) | 27.68 (2.67) | 29.75 (2.38) | 27.97 (2.0) | 25.24 (2.01) |
| FR.AO | 23.82 (1.52) | 29.23 (2.26) | 39.81 (3.83) | 35.53 (2.97) | 25.28 (2.06) | 28.45 (2.07) | 27.2 (1.81) | 22.8 (1.64) |
| FR.AR | 23.03 (1.6) | 30.95 (2.15) | 43.67 (4.16) | 39.63 (3.13) | 24.97 (1.94) | 29.7 (2.17) | 28.66 (2.04) | 24.16 (1.69) |
| FR.BF | 23.92 (1.47) | 37.13 (2.89) | 43.82 (4.52) | 40.64 (4.0) | 26.45 (2.04) | 31.58 (2.43) | 29.69 (1.99) | 24.23 (1.56) |
| FR.BT | 23.93 (1.52) | 31.98 (2.59) | 37.9 (4.14) | 35.58 (3.51) | 26.37 (2.53) | 28.15 (2.35) | 27.25 (2.22) | 25.05 (2.0) |
| FR.CE | 30.93 (3.06) | 97.99 (42.98) | 45.09 (4.9) | 39.91 (3.04) | 28.02 (2.27) | 43.79 (9.26) | 35.28 (2.67) | 32.07 (3.83) |
| FR.CN | 23.18 (1.48) | 36.75 (2.94) | 42.6 (5.77) | 38.22 (5.37) | 27.33 (4.09) | 30.64 (3.01) | 28.2 (2.01) | 24.26 (1.63) |
| FR.IF | 20.71 (1.3) | 29.75 (2.11) | 35.7 (2.97) | 31.07 (2.33) | 21.84 (2.06) | 24.52 (1.84) | 25.15 (1.96) | 20.05 (1.53) |
| FR.LP | 22.65 (1.4) | 30.01 (2.18) | 43.24 (4.8) | 38.56 (3.79) | 24.71 (2.34) | 28.87 (2.42) | 27.44 (2.14) | 23.38 (1.74) |
| FR.NC | 23.16 (1.49) | 28.39 (2.09) | 44.57 (5.77) | 38.66 (4.11) | 23.51 (1.98) | 28.7 (2.55) | 26.8 (2.0) | 23.86 (2.12) |
| FR.ND | 22.96 (1.38) | 32.37 (2.36) | 37.78 (3.32) | 33.16 (2.59) | 24.53 (2.15) | 26.6 (2.07) | 25.96 (2.05) | 21.79 (1.43) |
| FR.PL | 25.05 (1.7) | 33.53 (2.59) | 45.56 (6.55) | 41.66 (5.85) | 26.64 (2.72) | 31.14 (3.21) | 28.78 (2.46) | 26.55 (2.41) |
| FR.PR | 20.14 (1.32) | 30.12 (2.24) | 37.77 (3.3) | 34.09 (2.47) | 21.82 (1.62) | 26.01 (1.86) | 25.56 (1.89) | 20.59 (1.48) |

**Table S3.1.4:** Base models versus baseline ensemble methods for COVID-19 cases in France. The performances are given as the mean MAPE and its standard error in parentheses. The regions are given in HASC Codes (http://www.statoids.com/ihasc.html).

| Geography | LR | LSTM | XG | RF | ARIMA | Mean | Median | Prev.-Best |
|---|---|---|---|---|---|---|---|---|
| FR | 16.45 (1.01) | 33.61 (1.94) | 25.95 (1.66) | 24.26 (1.43) | 16.84 (1.05) | 20.43 (1.19) | 21.02 (1.22) | 17.57 (1.11) |
| FR.AC | 27.27 (1.46) | 26.43 (1.45) | 29.44 (1.8) | 27.52 (1.68) | 24.1 (1.4) | 24.32 (1.33) | 25.34 (1.42) | 23.5 (1.42) |
| FR.AO | 25.34 (2.0) | 26.66 (1.56) | 25.09 (1.62) | 23.33 (1.43) | 21.48 (1.36) | 21.48 (1.37) | 22.15 (1.37) | 20.66 (1.26) |
| FR.AR | 23.62 (3.36) | 28.65 (1.78) | 27.74 (2.26) | 26.34 (1.92) | 21.73 (1.67) | 22.73 (1.93) | 23.48 (1.71) | 23.2 (3.33) |
| FR.BF | 28.99 (4.16) | 27.4 (1.48) | 27.35 (1.85) | 25.06 (1.53) | 23.59 (1.34) | 23.42 (1.52) | 23.54 (1.29) | 22.64 (1.34) |
| FR.BT | 27.55 (2.13) | 28.7 (1.89) | 29.24 (2.07) | 26.89 (1.84) | 22.98 (1.36) | 23.79 (1.42) | 25.45 (1.58) | 24.84 (2.02) |
| FR.CE | 47.12 (5.49) | 41.03 (5.01) | 34.56 (2.93) | 30.39 (2.24) | 29.47 (2.15) | 31.7 (2.3) | 32.39 (1.71) | 39.67 (5.34) |
| FR.CN | 34.41 (3.86) | 32.32 (2.37) | 33.6 (2.5) | 30.9 (2.03) | 26.98 (1.85) | 28.01 (2.05) | 28.83 (1.88) | 26.45 (1.75) |
| FR.IF | 22.91 (1.71) | 28.58 (1.54) | 27.27 (1.88) | 25.55 (1.58) | 21.09 (1.31) | 22.55 (1.36) | 23.3 (1.38) | 22.32 (1.72) |
| FR.LP | 25.56 (2.83) | 26.26 (1.75) | 28.48 (1.85) | 26.71 (1.71) | 23.58 (1.53) | 23.27 (1.5) | 25.18 (1.62) | 20.98 (1.25) |
| FR.NC | 25.89 (2.58) | 26.89 (1.47) | 28.03 (2.01) | 25.21 (1.57) | 23.74 (1.39) | 23.15 (1.4) | 23.63 (1.38) | 24.45 (2.32) |
| FR.ND | 29.23 (1.87) | 31.09 (2.44) | 30.76 (1.9) | 27.35 (1.61) | 25.8 (1.43) | 24.7 (1.46) | 25.69 (1.43) | 26.2 (1.61) |
| FR.PL | 22.93 (1.47) | 28.35 (1.95) | 28.12 (2.0) | 25.96 (1.67) | 22.56 (1.41) | 22.53 (1.4) | 23.39 (1.48) | 22.85 (1.45) |
| FR.PR | 19.34 (1.19) | 25.51 (1.61) | 28.54 (2.0) | 25.81 (1.52) | 19.21 (1.16) | 21.54 (1.23) | 22.49 (1.28) | 19.28 (1.1) |

**Table S3.1.5:** Base models versus baseline ensemble methods for COVID-19 hospitalization in France. The performances are given as the mean MAPE and its standard error in parentheses. The regions are given in HASC Codes (http://www.statoids.com/ihasc.html).

| Geography | LR | LSTM | XG | RF | ARIMA | Mean | Median | Prev.-Best |
|---|---|---|---|---|---|---|---|---|
| FR | 16.77 (1.35) | 30.63 (1.87) | 20.79 (1.27) | 19.32 (1.19) | 17.09 (1.12) | 17.07 (1.03) | 17.96 (1.10) | 16.70 (1.42) |
| FR.AC | 36.32 (3.09) | 21.39 (1.29) | 27.0 (1.83) | 24.52 (1.42) | 22.7 (1.41) | 24.14 (1.33) | 24.92 (1.28) | 25.0 (1.65) |
| FR.AO | 31.27 (2.05) | 19.94 (1.14) | 24.87 (1.77) | 23.0 (1.51) | 21.77 (1.23) | 21.43 (1.13) | 22.42 (1.17) | 22.87 (1.39) |
| FR.AR | 28.14 (1.61) | 22.71 (1.48) | 26.66 (1.52) | 24.7 (1.46) | 21.55 (1.22) | 21.68 (1.26) | 23.0 (1.4) | 22.28 (1.34) |
| FR.BF | 37.52 (3.51) | 20.13 (1.2) | 26.11 (1.61) | 23.41 (1.29) | 21.63 (1.05) | 24.05 (1.26) | 25.71 (1.24) | 21.99 (1.2) |
| FR.BT | 35.44 (1.75) | 22.99 (2.16) | 27.96 (1.92) | 25.55 (1.45) | 23.57 (1.32) | 25.39 (1.2) | 27.52 (1.27) | 24.32 (1.39) |
| FR.CE | 46.49 (1.54) | 22.55 (5.19) | 21.96 (2.18) | 21.05 (1.78) | 18.39 (1.45) | 32.78 (1.19) | 40.17 (1.16) | 23.15 (5.13) |
| FR.CN | 36.17 (1.87) | 22.12 (1.59) | 28.53 (2.14) | 25.19 (1.28) | 23.49 (1.31) | 23.51 (1.16) | 25.68 (1.24) | 22.81 (1.29) |
| FR.IF | 27.36 (4.01) | 25.56 (1.71) | 24.24 (1.71) | 22.3 (1.48) | 21.38 (1.41) | 20.83 (1.54) | 21.06 (1.31) | 23.3 (3.98) |
| FR.LP | 30.95 (1.72) | 23.38 (1.47) | 27.25 (1.74) | 25.96 (1.63) | 23.99 (1.34) | 23.99 (1.27) | 25.79 (1.36) | 22.29 (1.38) |
| FR.NC | 37.14 (9.0) | 20.58 (1.19) | 25.51 (2.16) | 23.4 (1.68) | 21.94 (1.49) | 22.79 (2.36) | 21.52 (1.24) | 22.21 (1.16) |
| FR.ND | 30.01 (1.54) | 21.44 (1.45) | 26.39 (1.65) | 22.82 (1.17) | 21.35 (1.05) | 22.35 (1.03) | 24.65 (1.14) | 22.76 (1.32) |
| FR.PL | 32.14 (1.66) | 22.12 (2.04) | 24.14 (1.4) | 22.48 (1.25) | 20.23 (1.07) | 23.4 (1.08) | 25.71 (1.19) | 23.04 (1.34) |
| FR.PR | 27.82 (1.89) | 22.21 (1.38) | 26.14 (1.67) | 22.98 (1.33) | 20.86 (1.35) | 21.69 (1.24) | 22.63 (1.3) | 21.96 (1.47) |

**Table S3.1.6:** Base models versus baseline ensemble methods for COVID-19 deaths in France. The performances are given as the mean MAPE and its standard error in parentheses. The regions are given in HASC Codes (http://www.statoids.com/ihasc.html).

##### 3.2 Baseline Ensembles versus Dynamic Model Stacking and Selection

| Geography | LR | LSTM | XG | RF | ARIMA | Mean | Median | Prev.-Best | Selection | Stacking |
|---|---|---|---|---|---|---|---|---|---|---|
| **Daily COVID-19 Cases DE (N=28)** | | | | | | | | | | |
| DE | 19.04 (3.66) | 31.45 (4.05) | 34.37 (4.96) | 26.77 (3.23) | 23.63 (3.04) | 23.75 (2.51) | 24.40 (2.89) | 17.07 (2.38) | 28.94 (7.10) | 24.58 (2.41) |
| DE_reg | 24.36 (3.42) | 28.84 (2.78) | 34.04 (4.22) | 29.31 (3.01) | 25.55 (2.89) | 25.58 (2.68) | 26.53 (2.80) | 22.50 (2.13) | 30.44 (3.59) | 26.29 (2.37) |
| **Daily COVID-19 Hospitalization DE (N=28)** | | | | | | | | | | |
| DE | 15.80 (2.36) | 32.73 (4.36) | 25.37 (3.26) | 24.63 (2.88) | 9.77 (1.23) | 18.84 (2.26) | 19.37 (2.68) | 12.25 (2.01) | 24.51 (3.38) | 14.32 (2.01) |
| DE_reg | 21.63 (1.93) | 24.3 (2.01) | 31.03 (3.40) | 27.85 (2.84) | 14.28 (1.24) | 20.88 (2.05) | 21.04 (1.77) | 17.47 (1.36) | 24.39 (2.16) | 19.68 (2.05) |
| **Daily COVID-19 Deaths DE (N=28)** | | | | | | | | | | |
| DE | 22.89 (3.34) | 41.30 (4.66) | 37.85 (5.64) | 32.63 (3.99) | 24.56 (2.96) | 25.98 (3.05) | 26.86 (3.15) | 24.87 (3.83) | 27.27 (4.57) | 21.99 (2.40) |
| DE_reg | 37.80 (2.49) | 33.36 (4.96) | 39.46 (3.98) | 32.81 (2.53) | 28.79 (2.28) | 25.68 (1.17) | 26.63 (1.28) | 31.71 (1.78) | 34.59 (8.26) | 22.86 (2.70) |
| **Daily COVID-19 Cases FR (N=28)** | | | | | | | | | | |
| FR | 23.08 (2.50) | 37.84 (6.45) | 35.63 (6.79) | 32.28 (5.50) | 20.62 (3.36) | 25.13 (4.82) | 27.30 (5.12) | 21.53 (3.08) | 31.36 (8.19) | 20.48 (2.58) |
| FR_reg | 24.73 (2.43) | 30.0 (5.45) | 35.72 (6.22) | 33.26 (4.85) | 22.32 (3.46) | 26.2 (4.16) | 26.75 (3.95) | 22.80 (2.90) | 28.79 (5.23) | 22.09 (2.46) |
| **Daily COVID-19 Hospitalization FR (N=28)** | | | | | | | | | | |
| FR | 19.82 (2.08) | 39.23 (4.39) | 31.82 (3.71) | 27.27 (3.05) | 19.11 (1.92) | 23.36 (2.60) | 24.79 (2.62) | 21.35 (2.76) | 29.80 (5.04) | 17.60 (2.62) |
| FR_reg | 28.09 (1.73) | 35.15 (3.59) | 34.75 (3.46) | 30.55 (2.50) | 26.16 (1.85) | 26.65 (1.99) | 27.28 (2.12) | 28.88 (2.09) | 32.53 (4.03) | 20.73 (1.82) |
| **Daily COVID-19 Deaths FR (N=28)** | | | | | | | | | | |
| FR | 18.20 (3.34) | 34.05 (4.66) | 22.82 (5.64) | 19.75 (3.99) | 19.43 (2.96) | 18.82 (3.05) | 18.61 (3.15) | 19.32 (3.83) | 21.06 (4.57) | 14.07 (2.40) |
| FR_reg | 34.72 (2.49) | 24.83 (4.96) | 30.26 (3.98) | 26.47 (2.53) | 23.24 (2.28) | 23.09 (1.17) | 24.41 (1.28) | 26.76 (1.78) | 24.41 (8.26) | 17.99 (2.70) |
| **Weekly Influenza Cases DE (N=6)** | | | | | | | | | | |
| DE | 48.50 (1.08) | 4.20 (1.19) | 2.67 (0.58 | 4.86 (1.47) | 2.82 (1.49) | 36.51 (1.42) | 45.15 (1.58) | 10.87 (7.76) | 35.80 (8.17) | 7.82 (3.82) |
| **Weekly SARI Hospitalization DE (N=16)** | | | | | | | | | | |
| DE | 19.87 (3.64) | 16.66 (3.0) | 13.61 (3.74) | 14.21 (3.79) | 12.54 (3.45) | 12.89 (2.57) | 14.02 (3.26) | 15.15 (3.66) | 17.2 (2.64) | 13.19 (3.12) |
| **Consensus Ranking** | | | | | | | | | | |
| All | 6 | 7 | 9 | 8 | 2 | 4 | 5 | 2 | 3 | 1 |

| Best Model | 2nd Best Model | 3rd Best Model |
|---|---|---|

**Table S3.2.1:** Ensemble Model Pipeline Results. The performances are given as the mean MAPE and its standard error in parentheses of the N test windows for each dataset / dataset aggregation. The best three models are colored according to the provided legend. DE (FR) stands for German (France) country level and DE_reg (FR_reg) for German (France) regional level aggregated to country level.

| | ARIMA | LR | LSTM | Mean | Median | Prev.-Best | RF | Selection | Stacking |
|---|---|---|---|---|---|---|---|---|---|
| **LR** | 1.46E-15 | - | - | - | - | - | - | - | - |
| **LSTM** | <1E-16 | 2.79E-01 | - | - | - | - | - | - | - |
| **Mean** | 3.52E-04 | 9.54E-05 | 8.24E-10 | - | - | - | - | - | - |
| **Median** | 1.88E-08 | 6.27E-02 | 4.66E-05 | 2.79E-01 | - | - | - | - | - |
| **Prev.-Best** | 1.94E-01 | 1.38E-08 | 4.23E-15 | 2.79E-01 | 1.95E-03 | - | - | - | - |
| **RF** | <1E-16 | 4.97E-03 | 3.01E-01 | 4.14E-15 | 4.54E-09 | <1E-16 | - | - | - |
| **Selection** | 4.97E-03 | 1.01E-04 | 1.09E-09 | 6.30E-01 | 1.94E-01 | 3.24E-01 | 1.87E-14 | - | - |
| **Stacking** | 5.74E-03 | <1E-16 | <1E-16 | 1.01E-13 | <1E-16 | 2.57E-07 | <1E-16 | 2.17E-10 | - |
| **XG** | <1E-16 | 1.17E-10 | 6.91E-06 | <1E-16 | <1E-16 | <1E-16 | 4.27E-03 | <1E-16 | <1E-16 |

**Table S3.2.2:** Base models versus baseline ensembles versus Dynamic Model Stacking and Selection approaches: pairwise Wilcox Test (adjusted p-values).

| Geography | LR | LSTM | XG | RF | ARIMA | Mean | Median | Prev.-Best | Selection | Stacking |
|---|---|---|---|---|---|---|---|---|---|---|
| DE | 19.04 (1.64) | 31.45 (1.81) | 34.37 (2.22) | 26.77 (1.45) | 23.63 (1.36) | 23.75 (1.12) | 24.4 (1.29) | 17.07 (1.07) | 28.94 (7.1) | 24.58 (2.41) |
| DE.BB | 22.84 (1.53) | 24.65 (1.47) | 31.78 (2.27) | 27.87 (1.79) | 25.02 (1.73) | 23.73 (1.58) | 25.13 (1.62) | 22.08 (1.1) | 31.01 (5.59) | 26.39 (3.68) |
| DE.BE | 27.51 (2.88) | 24.48 (1.15) | 26.61 (1.78) | 25.34 (1.56) | 24.48 (1.28) | 23.44 (1.33) | 22.76 (1.3) | 19.32 (1.11) | 25.33 (6.48) | 19.98 (2.6) |
| DE.BW | 25.49 (1.61) | 29.83 (2.0) | 41.49 (3.05) | 36.51 (2.6) | 29.53 (2.09) | 29.16 (1.94) | 30.74 (2.01) | 26.37 (1.73) | 29.4 (3.77) | 30.99 (3.23) |
| DE.BY | 26.52 (2.28) | 31.54 (2.11) | 38.93 (2.71) | 35.03 (2.2) | 28.16 (1.78) | 29.34 (1.66) | 30.49 (1.85) | 26.81 (1.92) | 27.74 (4.23) | 27.97 (3.14) |
| DE.HB | 25.12 (2.16) | 43.76 (3.58) | 36.96 (3.16) | 28.16 (1.64) | 25.38 (1.43) | 28.81 (1.62) | 27.73 (1.56) | 22.52 (1.1) | 43.13 (9.22) | 30.99 (3.24) |
| DE.HE | 20.03 (1.61) | 27.37 (1.6) | 32.43 (1.91) | 29.1 (1.48) | 24.63 (1.38) | 24.64 (1.21) | 26.52 (1.29) | 18.95 (1.15) | 24.38 (3.27) | 24.19 (2.66) |
| DE.HH | 20.51 (1.33) | 28.69 (1.81) | 26.91 (1.97) | 23.29 (1.42) | 21.06 (1.61) | 21.75 (1.41) | 22.19 (1.48) | 20.55 (1.5) | 23.26 (3.39) | 21.76 (2.64) |
| DE.MV | 21.94 (1.68) | 24.02 (1.14) | 28.41 (1.97) | 22.36 (1.34) | 20.98 (1.34) | 21.2 (1.29) | 21.64 (1.27) | 18.92 (1.06) | 35.92 (5.56) | 28.9 (3.88) |
| DE.NI | 22.23 (2.05) | 24.0 (1.37) | 30.06 (1.73) | 27.58 (1.26) | 24.13 (1.27) | 22.34 (1.16) | 24.54 (1.3) | 20.09 (1.17) | 28.58 (4.57) | 24.17 (2.83) |
| DE.NW | 19.06 (1.53) | 20.66 (1.52) | 26.0 (1.67) | 21.38 (1.36) | 21.48 (1.21) | 18.28 (1.14) | 20.0 (1.17) | 19.45 (1.47) | 25.28 (4.11) | 22.64 (2.4) |
| DE.RP | 30.86 (2.66) | 32.39 (1.81) | 39.39 (2.71) | 34.31 (2.25) | 30.74 (2.2) | 30.19 (2.1) | 30.69 (2.25) | 26.17 (2.15) | 26.7 (3.89) | 25.45 (3.16) |
| DE.SH | 22.43 (1.96) | 22.96 (1.3) | 30.04 (1.93) | 26.78 (1.36) | 25.7 (1.5) | 22.89 (1.33) | 25.45 (1.44) | 20.82 (1.86) | 31.46 (4.65) | 26.85 (3.02) |
| DE.SL | 29.98 (3.08) | 40.37 (2.48) | 49.73 (4.07) | 41.86 (3.14) | 30.66 (2.62) | 35.1 (2.55) | 35.26 (2.68) | 23.77 (1.58) | 57.54 (18.31) | 36.84 (6.57) |
| DE.SN | 23.17 (1.83) | 26.02 (1.41) | 33.39 (2.12) | 29.4 (1.53) | 24.68 (1.61) | 24.78 (1.42) | 25.89 (1.46) | 22.22 (1.36) | 25.82 (4.54) | 23.72 (3.1) |
| DE.ST | 27.81 (2.11) | 28.36 (1.59) | 34.65 (2.31) | 29.61 (1.88) | 26.5 (1.6) | 26.72 (1.65) | 28.03 (1.64) | 26.04 (1.7) | 24.41 (3.71) | 24.76 (3.6) |
| DE.TH | 24.3 (1.87) | 32.31 (1.69) | 37.79 (2.88) | 30.42 (1.95) | 25.61 (1.61) | 26.94 (1.64) | 27.49 (1.63) | 25.88 (1.96) | 27.05 (5.1) | 25.0 (3.76) |

**Table S3.2.3:** Base models versus baseline ensemble methods versus Dynamic Stacking and Selection for COVID-19 cases in Germany. The performances are given as the mean MAPE and its standard error in parentheses. The regions are given in HASC Codes (http://www.statoids.com/ihasc.html).

| Geography | LR | LSTM | XG | RF | ARIMA | Mean | Median | Prev.-Best | Selection | Stacking |
|---|---|---|---|---|---|---|---|---|---|---|
| DE | 15.8 (1.06) | 32.73 (1.96) | 25.37 (1.47) | 24.63 (1.3) | 9.77 (0.55) | 18.84 (1.02) | 19.37 (1.21) | 12.25 (0.91) | 24.51 (3.83) | 14.32 (2.01) |
| DE.BB | 20.25 (1.15) | 20.52 (1.18) | 31.42 (1.72) | 26.98 (1.7) | 12.72 (0.76) | 19.85 (1.15) | 19.58 (1.11) | 13.49 (0.72) | 33.5 (5.31) | 22.5 (3.23) |
| DE.BE | 20.26 (1.06) | 17.04 (1.36) | 25.41 (1.39) | 22.9 (1.31) | 11.91 (0.85) | 16.66 (1.05) | 16.11 (1.08) | 16.01 (1.05) | 18.93 (3.7) | 15.11 (2.36) |
| DE.BW | 19.36 (1.33) | 24.69 (1.19) | 32.17 (1.95) | 28.34 (1.92) | 12.94 (0.7) | 20.66 (1.3) | 22.2 (1.29) | 15.74 (0.96) | 21.83 (2.29) | 15.25 (1.77) |
| DE.BY | 18.93 (1.39) | 24.99 (1.12) | 29.51 (2.16) | 26.29 (1.92) | 10.94 (0.68) | 19.37 (1.3) | 19.65 (1.24) | 14.01 (0.89) | 20.15 (3.01) | 15.27 (1.92) |
| DE.HB | 37.6 (2.64) | 43.87 (2.88) | 49.32 (3.86) | 44.51 (3.45) | 24.88 (1.87) | 34.88 (2.42) | 35.91 (2.4) | 33.31 (2.0) | 24.31 (3.1) | 25.99 (2.98) |
| DE.HE | 15.69 (1.23) | 22.95 (1.13) | 29.29 (2.03) | 27.51 (1.9) | 10.94 (0.62) | 19.47 (1.23) | 19.54 (1.12) | 11.19 (0.63) | 20.83 (3.77) | 15.04 (1.46) |
| DE.HH | 26.68 (1.78) | 28.9 (1.86) | 30.64 (2.14) | 28.79 (1.97) | 18.3 (1.22) | 23.07 (1.62) | 23.33 (1.68) | 20.97 (1.51) | 21.95 (2.73) | 21.22 (4.99) |
| DE.MV | 17.29 (1.13) | 23.25 (1.57) | 26.78 (1.96) | 23.08 (1.34) | 12.61 (0.78) | 17.71 (1.2) | 18.46 (1.18) | 14.4 (0.88) | 29.35 (4.57) | 28.87 (6.62) |
| DE.NI | 17.58 (1.01) | 20.6 (0.97) | 26.57 (1.56) | 24.49 (1.67) | 11.98 (0.69) | 18.33 (0.94) | 18.58 (0.9) | 12.08 (0.84) | 20.44 (3.45) | 15.78 (2.3) |
| DE.NW | 14.68 (0.98) | 21.5 (1.25) | 20.65 (1.33) | 20.44 (1.27) | 10.2 (0.6) | 16.3 (0.83) | 16.6 (0.92) | 12.58 (0.9) | 21.91 (3.27) | 12.3 (1.88) |
| DE.RP | 19.89 (1.72) | 23.54 (1.31) | 30.34 (2.08) | 25.88 (1.87) | 14.61 (0.97) | 19.83 (1.36) | 18.95 (1.38) | 16.61 (1.23) | 21.1 (3.5) | 15.89 (2.28) |
| DE.SH | 19.73 (0.92) | 19.61 (1.35) | 23.51 (1.18) | 21.49 (1.17) | 15.96 (1.07) | 16.75 (0.95) | 16.81 (0.9) | 16.56 (0.96) | 23.82 (4.2) | 19.09 (2.76) |
| DE.SL | 30.53 (1.85) | 28.27 (1.34) | 46.54 (3.6) | 42.85 (3.31) | 20.53 (1.16) | 28.49 (2.1) | 28.33 (1.7) | 26.45 (1.35) | 40.21 (6.97) | 35.86 (11.74) |
| DE.SN | 21.97 (1.34) | 23.37 (1.22) | 30.85 (2.0) | 27.19 (1.74) | 13.04 (0.85) | 20.63 (1.21) | 20.59 (1.09) | 17.91 (1.15) | 25.92 (3.26) | 19.88 (2.78) |
| DE.ST | 22.5 (1.65) | 22.09 (1.38) | 34.47 (2.45) | 28.84 (1.27) | 13.02 (0.74) | 22.14 (1.14) | 21.21 (1.07) | 15.39 (0.78) | 26.53 (3.32) | 21.03 (3.58) |
| DE.TH | 23.15 (1.06) | 23.55 (1.29) | 29.07 (1.51) | 26.09 (1.25) | 13.94 (0.93) | 19.89 (1.08) | 20.71 (1.08) | 22.75 (1.34) | 19.38 (2.62) | 15.75 (1.33) |

**Table S3.2.4:** Base models versus baseline ensemble methods versus Dynamic Stacking and Selection for COVID-19 hospitalization in Germany. The performances are given as the mean MAPE and its standard error in parentheses. The regions are given in HASC Codes (http://www.statoids.com/ihasc.html).

| Geography | LR | LSTM | XG | RF | ARIMA | Mean | Median | Prev.-Best | Selection | Stacking |
|---|---|---|---|---|---|---|---|---|---|---|
| DE | 22.89 (1.51) | 41.3 (2.1) | 37.85 (2.54) | 32.63 (1.8) | 24.56 (1.33) | 25.98 (1.37) | 26.86 (1.42) | 24.87 (1.72) | 27.27 (4.57) | 21.99 (2.4) |
| DE.BB | 31.56 (1.65) | 29.1 (3.29) | 35.55 (2.28) | 29.2 (1.61) | 26.49 (1.57) | 21.75 (1.02) | 21.08 (1.02) | 27.95 (1.54) | 32.73 (12.23) | 17.97 (2.98) |
| DE.BE | 47.34 (5.1) | 31.41 (2.0) | 42.25 (3.41) | 33.58 (2.6) | 30.33 (2.18) | 26.13 (2.12) | 26.97 (1.52) | 33.41 (2.2) | 39.17 (13.46) | 22.57 (5.5) |
| DE.BW | 31.83 (2.03) | 30.89 (1.59) | 44.74 (2.99) | 38.67 (2.15) | 30.49 (1.75) | 27.33 (1.44) | 28.61 (1.51) | 36.22 (2.34) | 30.33 (4.94) | 23.97 (3.75) |
| DE.BY | 35.03 (1.8) | 32.45 (1.67) | 41.84 (2.79) | 35.86 (2.46) | 29.74 (1.37) | 27.79 (1.57) | 28.75 (1.53) | 33.28 (2.01) | 35.18 (5.7) | 25.0 (2.9) |
| DE.HB | 39.88 (1.32) | 41.62 (5.47) | 27.5 (1.81) | 23.41 (1.09) | 19.86 (0.85) | 25.93 (0.99) | 33.02 (1.11) | 22.08 (1.18) | 42.29 (21.99) | 40.06 (22.01) |
| DE.HE | 34.43 (3.44) | 31.5 (2.31) | 44.97 (3.33) | 34.39 (2.48) | 31.53 (2.07) | 26.88 (1.58) | 25.92 (1.64) | 36.48 (3.41) | 26.28 (4.66) | 21.67 (2.61) |
| DE.HH | 41.6 (2.76) | 33.14 (3.0) | 41.7 (1.73) | 33.44 (1.48) | 27.62 (1.24) | 23.47 (1.07) | 23.43 (1.15) | 33.41 (1.7) | 33.51 (9.38) | 18.01 (2.29) |
| DE.MV | 38.46 (1.52) | 34.83 (4.63) | 34.35 (2.16) | 31.04 (1.75) | 27.44 (1.57) | 23.92 (1.16) | 25.38 (1.18) | 27.37 (1.74) | 36.69 (13.73) | 16.52 (1.86) |
| DE.NI | 25.99 (1.42) | 30.13 (1.38) | 40.55 (2.35) | 36.73 (1.93) | 28.83 (1.74) | 25.69 (1.38) | 27.65 (1.46) | 25.75 (0.92) | 26.95 (4.24) | 24.35 (2.86) |
| DE.NW | 26.69 (1.55) | 31.23 (1.86) | 32.16 (2.29) | 29.26 (1.72) | 27.1 (1.72) | 24.78 (1.33) | 25.83 (1.45) | 27.97 (1.56) | 24.14 (3.5) | 23.02 (2.8) |
| DE.RP | 43.54 (2.48) | 34.68 (2.2) | 51.48 (5.04) | 39.73 (2.6) | 37.0 (2.45) | 31.45 (1.86) | 31.55 (1.57) | 38.29 (2.44) | 37.63 (10.46) | 23.05 (3.23) |
| DE.SH | 33.2 (1.77) | 29.81 (3.5) | 33.04 (1.81) | 27.57 (1.31) | 25.41 (1.41) | 21.05 (0.98) | 21.29 (1.02) | 27.53 (1.31) | 35.71 (11.34) | 19.36 (2.42) |
| DE.SL | 43.55 (2.12) | 37.24 (3.47) | 36.64 (1.65) | 31.7 (1.49) | 29.38 (1.24) | 24.83 (1.18) | 27.39 (1.35) | 39.45 (2.38) | 52.87 (19.98) | 29.35 (7.34) |
| DE.SN | 45.04 (2.21) | 33.82 (3.24) | 44.3 (2.79) | 33.52 (2.14) | 31.6 (1.9) | 27.37 (1.3) | 25.93 (1.35) | 31.22 (1.99) | 35.55 (5.54) | 25.23 (3.19) |
| DE.ST | 48.73 (4.48) | 37.95 (3.18) | 42.06 (2.72) | 35.48 (1.92) | 30.82 (1.41) | 29.35 (1.27) | 29.01 (1.08) | 38.54 (4.14) | 31.8 (9.82) | 18.1 (2.58) |
| DE.TH | 37.97 (3.35) | 34.02 (3.45) | 38.19 (2.79) | 31.37 (2.17) | 27.04 (2.31) | 23.19 (1.41) | 24.19 (1.37) | 28.47 (1.51) | 32.63 (8.87) | 17.47 (1.95) |

**Table S3.2.5:** Base models versus baseline ensemble methods versus Dynamic Stacking and Selection for COVID-19 deaths in Germany. The performances are given as the mean MAPE and its standard error in parentheses. The regions are given in HASC Codes (http://www.statoids.com/ihasc.html).

| Geography | LR | LSTM | XG | RF | ARIMA | Mean | Median | Prev.-Best | Selection | Stacking |
|---|---|---|---|---|---|---|---|---|---|---|
| FR | 23.08 (1.13) | 37.84 (2.9) | 35.63 (3.06) | 32.28 (2.48) | 20.62 (1.51) | 25.13 (2.17) | 27.3 (2.31) | 21.53 (1.39) | 31.36 (8.19) | 20.48 (2.58) |
| FR.AC | 22.86 (1.37) | 28.36 (2.16) | 35.61 (3.18) | 32.44 (2.74) | 22.8 (1.76) | 25.91 (1.96) | 27.18 (2.05) | 25.48 (1.85) | 21.91 (3.66) | 23.27 (3.09) |
| FR.AO | 27.57 (1.59) | 31.28 (3.14) | 37.88 (2.99) | 31.9 (2.29) | 27.05 (2.86) | 28.43 (2.22) | 28.88 (1.98) | 21.69 (1.71) | 21.35 (3.21) | 22.31 (2.94) |
| FR.AR | 22.1 (1.22) | 27.71 (2.02) | 37.92 (3.12) | 35.87 (2.62) | 21.29 (1.56) | 25.75 (1.93) | 26.69 (1.72) | 22.77 (2.08) | 31.07 (11.45) | 21.48 (2.7) |
| FR.BF | 24.82 (1.31) | 34.58 (3.4) | 36.91 (2.99) | 34.3 (2.62) | 22.87 (1.79) | 28.09 (2.18) | 28.12 (2.0) | 23.69 (1.31) | 33.82 (9.05) | 23.61 (2.81) |
| FR.BT | 26.24 (1.77) | 27.8 (2.66) | 32.46 (3.13) | 31.47 (2.67) | 22.87 (2.0) | 25.35 (2.27) | 26.76 (2.4) | 24.28 (2.48) | 24.49 (3.54) | 24.15 (3.16) |
| FR.CE | 28.57 (1.61) | 34.59 (2.94) | 30.94 (1.87) | 30.42 (1.63) | 24.61 (1.16) | 27.03 (1.42) | 26.8 (1.3) | 21.35 (1.25) | 46.22 (18.99) | 20.95 (2.68) |
| FR.CN | 25.53 (1.6) | 37.25 (3.86) | 38.57 (3.19) | 32.51 (2.14) | 22.08 (1.91) | 27.95 (2.36) | 28.7 (1.96) | 25.29 (1.83) | 40.39 (18.19) | 23.8 (2.87) |
| FR.IF | 23.16 (1.17) | 28.82 (1.62) | 31.75 (2.56) | 30.18 (2.06) | 17.07 (1.23) | 22.3 (1.58) | 22.89 (1.47) | 20.28 (1.53) | 25.1 (7.38) | 18.26 (2.66) |
| FR.LP | 23.82 (1.17) | 27.07 (2.34) | 37.3 (3.79) | 35.46 (2.85) | 22.77 (1.63) | 25.77 (2.2) | 26.5 (2.24) | 19.31 (1.04) | 31.89 (8.31) | 25.28 (3.05) |
| FR.NC | 23.45 (1.26) | 28.08 (2.26) | 39.53 (2.87) | 36.54 (2.24) | 19.24 (1.31) | 26.54 (1.74) | 27.35 (1.78) | 23.51 (1.8) | 21.82 (2.96) | 19.17 (2.7) |
| FR.ND | 24.79 (1.31) | 30.63 (3.12) | 31.05 (2.45) | 30.32 (2.22) | 20.94 (1.56) | 24.31 (1.97) | 24.11 (1.77) | 20.37 (1.19) | 22.31 (2.55) | 21.47 (2.41) |
| FR.PL | 27.68 (1.63) | 30.42 (3.15) | 41.9 (3.74) | 37.66 (3.07) | 25.7 (2.1) | 29.86 (2.49) | 30.33 (2.76) | 29.28 (2.89) | 31.82 (6.5) | 23.97 (3.62) |
| FR.PR | 20.86 (0.92) | 23.34 (1.99) | 32.6 (3.08) | 33.33 (2.87) | 20.83 (1.42) | 23.33 (1.87) | 23.41 (1.79) | 19.15 (1.0) | 22.08 (4.36) | 19.49 (2.99) |

**Table S3.2.6:** Base models versus baseline ensemble methods versus Dynamic Stacking and Selection for COVID-19 cases in France. The performances are given as the mean MAPE and its standard error in parentheses. The regions are given in HASC Codes (http://www.statoids.com/ihasc.html).

| Geography | LR | LSTM | XG | RF | ARIMA | Mean | Median | Prev.-Best | Selection | Stacking |
|---|---|---|---|---|---|---|---|---|---|---|
| FR | 19.82 (0.94) | 39.23 (1.98) | 31.82 (1.67) | 27.27 (1.37) | 19.11 (0.87) | 23.36 (1.17) | 24.79 (1.18) | 21.35 (1.24) | 29.8 (5.04) | 17.6 (2.62) |
| FR.AC | 26.93 (1.11) | 29.22 (1.57) | 30.47 (1.8) | 27.46 (1.55) | 25.73 (1.47) | 25.24 (1.35) | 26.86 (1.39) | 26.62 (1.37) | 31.44 (3.79) | 22.11 (2.59) |
| FR.AO | 31.33 (2.15) | 33.94 (1.56) | 34.6 (1.98) | 29.67 (1.63) | 25.2 (1.27) | 27.52 (1.48) | 27.71 (1.61) | 26.16 (1.28) | 29.16 (5.45) | 19.07 (3.24) |
| FR.AR | 21.28 (0.86) | 29.06 (1.52) | 30.2 (1.79) | 26.16 (1.48) | 22.55 (1.2) | 21.48 (1.26) | 22.67 (1.27) | 21.87 (1.15) | 44.8 (15.82) | 27.44 (7.47) |
| FR.BF | 27.77 (1.58) | 37.09 (1.93) | 33.8 (1.68) | 29.88 (1.32) | 27.85 (1.28) | 27.21 (1.36) | 28.27 (1.36) | 29.35 (1.22) | 25.18 (3.23) | 16.78 (2.13) |
| FR.BT | 22.95 (1.26) | 31.91 (1.45) | 29.17 (1.5) | 24.07 (1.09) | 23.12 (0.97) | 22.44 (1.06) | 22.7 (1.04) | 24.79 (1.16) | 32.33 (5.95) | 19.18 (2.71) |
| FR.CE | 49.35 (4.19) | 56.3 (8.82) | 37.98 (2.66) | 34.32 (2.2) | 32.88 (2.29) | 31.1 (2.07) | 28.63 (1.52) | 57.67 (9.35) | 37.55 (6.77) | 22.0 (3.69) |
| FR.CN | 27.52 (1.81) | 45.15 (2.77) | 43.26 (3.05) | 37.56 (2.57) | 32.71 (2.05) | 32.81 (2.01) | 34.26 (2.22) | 29.7 (1.6) | 51.9 (16.04) | 27.35 (5.58) |
| FR.IF | 24.74 (1.19) | 32.18 (1.84) | 32.44 (1.96) | 29.46 (1.55) | 23.21 (1.55) | 24.91 (1.46) | 26.33 (1.51) | 25.97 (1.29) | 30.03 (7.64) | 20.14 (3.41) |
| FR.LP | 24.94 (1.59) | 25.22 (1.72) | 32.0 (2.19) | 27.87 (1.81) | 24.6 (1.46) | 24.06 (1.56) | 25.42 (1.59) | 24.65 (1.33) | 25.03 (2.95) | 18.22 (2.44) |
| FR.NC | 32.06 (2.71) | 33.64 (1.85) | 38.17 (2.22) | 31.11 (1.71) | 26.4 (1.6) | 29.04 (1.73) | 28.57 (1.63) | 27.83 (1.77) | 33.01 (6.47) | 20.32 (3.1) |
| FR.ND | 27.76 (1.49) | 38.9 (2.09) | 38.79 (2.35) | 35.5 (1.97) | 29.54 (1.49) | 29.11 (1.57) | 30.34 (1.56) | 30.7 (1.71) | 32.27 (4.47) | 21.51 (2.97) |
| FR.PL | 28.19 (2.08) | 39.62 (2.86) | 36.85 (2.87) | 33.1 (2.32) | 26.45 (1.9) | 28.03 (1.97) | 29.2 (2.13) | 30.52 (2.17) | 26.19 (3.53) | 18.28 (2.23) |
| FR.PR | 20.36 (1.05) | 24.77 (1.53) | 34.01 (2.15) | 31.05 (1.87) | 19.89 (0.92) | 23.46 (1.21) | 23.72 (1.3) | 19.59 (0.96) | 23.95 (3.25) | 17.07 (2.09) |

**Table S3.2.6:** Base models versus baseline ensemble methods versus Dynamic Stacking and Selection for COVID-19 hospitalization in France. The performances are given as the mean MAPE and its standard error in parentheses. The regions are given in HASC Codes (http://www.statoids.com/ihasc.html).

| Geography | LR | LSTM | XG | RF | ARIMA | Mean | Median | Prev.-Best | Selection | Stacking |
|---|---|---|---|---|---|---|---|---|---|---|
| FR | 18.2 (0.99) | 34.05 (2.05) | 22.82 (1.23) | 19.75 (1.1) | 19.43 (0.99) | 18.82 (1.05) | 18.61 (1.04) | 19.32 (1.57) | 21.06 (4.95) | 14.07 (2.17) |
| FR.AC | 31.28 (1.55) | 21.54 (1.18) | 29.78 (1.64) | 25.2 (1.11) | 23.3 (1.04) | 20.57 (0.92) | 20.77 (0.96) | 28.05 (1.58) | 20.01 (2.48) | 18.86 (2.28) |
| FR.AO | 30.23 (1.9) | 23.39 (1.52) | 30.44 (2.23) | 26.21 (1.89) | 22.35 (1.54) | 21.99 (1.23) | 21.63 (1.21) | 26.34 (1.83) | 32.1 (5.93) | 22.11 (2.93) |
| FR.AR | 28.83 (1.46) | 23.43 (1.22) | 28.63 (1.6) | 26.58 (1.41) | 21.44 (0.94) | 19.88 (0.9) | 19.84 (0.91) | 27.61 (1.28) | 26.16 (4.96) | 17.51 (2.32) |
| FR.BF | 37.07 (1.53) | 23.99 (1.25) | 28.17 (2.16) | 21.63 (1.23) | 20.06 (0.91) | 21.22 (0.97) | 22.04 (0.97) | 24.61 (1.52) | 18.4 (2.5) | 14.72 (1.64) |
| FR.BT | 42.02 (2.38) | 25.54 (1.66) | 38.2 (3.19) | 32.51 (2.17) | 30.21 (1.78) | 27.89 (1.32) | 30.13 (1.17) | 33.76 (1.87) | 22.77 (5.22) | 15.76 (1.93) |
| FR.CE | 43.29 (1.42) | 21.38 (2.4) | 19.1 (1.16) | 16.98 (0.85) | 15.55 (0.69) | 30.4 (1.0) | 40.05 (1.02) | 18.44 (1.24) | 27.3 (5.67) | 17.7 (5.11) |
| FR.CN | 42.54 (2.15) | 31.12 (2.25) | 34.5 (1.71) | 33.35 (1.37) | 27.6 (1.53) | 24.56 (1.29) | 24.86 (1.31) | 31.63 (1.56) | 21.47 (4.27) | 18.46 (2.75) |
| FR.IF | 29.9 (1.73) | 25.68 (1.75) | 31.41 (2.06) | 28.14 (1.74) | 25.36 (1.85) | 22.38 (1.28) | 22.6 (1.25) | 26.55 (1.8) | 38.71 (18.74) | 17.9 (2.27) |
| FR.LP | 29.88 (1.8) | 23.77 (1.55) | 29.72 (2.22) | 28.24 (1.92) | 23.07 (1.3) | 22.53 (1.2) | 23.69 (1.17) | 23.92 (1.4) | 20.84 (2.14) | 18.28 (1.59) |
| FR.NC | 37.4 (3.57) | 23.19 (1.36) | 27.61 (1.64) | 26.34 (1.63) | 22.79 (1.34) | 22.72 (1.54) | 21.58 (1.12) | 26.03 (1.37) | 27.17 (8.1) | 18.89 (2.46) |
| FR.ND | 27.8 (1.38) | 26.05 (2.34) | 31.1 (1.87) | 24.12 (1.3) | 20.67 (1.09) | 19.5 (0.79) | 22.22 (0.95) | 24.67 (1.44) | 21.09 (3.15) | 17.66 (2.64) |
| FR.PL | 36.77 (1.84) | 30.86 (2.29) | 32.71 (2.04) | 28.92 (1.71) | 26.13 (1.37) | 24.41 (1.09) | 26.3 (1.15) | 29.25 (1.72) | 20.58 (4.09) | 19.57 (4.1) |
| FR.PR | 34.34 (2.44) | 22.89 (1.14) | 31.98 (1.92) | 25.94 (1.48) | 23.57 (1.48) | 22.2 (1.24) | 21.65 (1.2) | 26.98 (1.42) | 20.74 (2.77) | 16.42 (2.15) |

**Table S3.2.6:** Base models versus baseline ensemble methods versus Dynamic Stacking and Selection for COVID-19 deaths in France. The performances are given as the mean MAPE and its standard error in parentheses. The regions are given in HASC Codes (http://www.statoids.com/ihasc.html).

### S3.3 Potential Benefits of Including Metadata

| Geography | Selection | Stacking | Sel. Meta | Stack. Meta |
|---|---|---|---|---|
| DE | 28.94 (7.1) | 24.58 (2.41) | 40.23 (8.68) | 22.26 (2.26) |
| DE.BB | 31.01 (5.59) | 26.39 (3.68) | 37.45 (7.08) | 26.38 (3.26) |
| DE.BE | 25.33 (6.48) | 19.98 (2.6) | 32.77 (6.5) | 19.45 (2.38) |
| DE.BW | 29.4 (3.77) | 30.99 (3.23) | 35.88 (6.36) | 30.7 (2.85) |
| DE.BY | 27.74 (4.23) | 27.97 (3.14) | 32.92 (5.48) | 27.8 (3.14) |
| DE.HB | 43.13 (9.22) | 30.99 (3.24) | 49.68 (9.28) | 26.74 (2.86) |
| DE.HE | 24.38 (3.27) | 24.19 (2.66) | 30.68 (5.09) | 24.06 (2.43) |
| DE.HH | 23.26 (3.39) | 21.76 (2.64) | 28.35 (4.1) | 19.57 (2.36) |
| DE.MV | 35.92 (5.56) | 28.9 (3.88) | 44.27 (8.04) | 27.85 (5.04) |
| DE.NI | 28.58 (4.57) | 24.17 (2.83) | 31.95 (5.3) | 22.97 (2.4) |
| DE.NW | 25.28 (4.11) | 22.64 (2.4) | 29.01 (4.99) | 20.97 (2.28) |
| DE.RP | 26.7 (3.89) | 25.45 (3.16) | 34.62 (5.16) | 24.73 (2.54) |
| DE.SH | 31.46 (4.65) | 26.85 (3.02) | 39.83 (6.29) | 26.82 (3.45) |
| DE.SL | 57.54 (18.31) | 36.84 (6.57) | 65.95 (18.62) | 35.64 (5.47) |
| DE.SN | 25.82 (4.54) | 23.72 (3.1) | 35.72 (5.39) | 24.76 (2.94) |
| DE.ST | 24.41 (3.71) | 24.76 (3.6) | 35.04 (6.14) | 24.73 (3.39) |
| DE.TH | 27.05 (5.1) | 25.0 (3.76) | 35.39 (6.72) | 25.29 (3.82) |

**Table S3.3.1:** Comparison of Dynamic Selection and Dynamic Stacking with and without the inclusion of metadata for COVID-19 Cases in Germany. The performances are given as the mean MAPE and its standard error in parentheses. The regions are given in HASC Codes (http://www.statoids.com/ihasc.html).

| Geography | Selection | Stacking | Sel. Meta | Stack. Meta |
|---|---|---|---|---|
| DE | 24.51 (3.83) | 14.32 (2.01) | 24.07 (7.19) | 14.49 (1.98) |
| DE.BB | 33.5 (5.31) | 22.5 (3.23) | 36.56 (8.09) | 22.12 (3.21) |
| DE.BE | 18.93 (3.7) | 15.11 (2.36) | 31.14 (9.04) | 14.81 (2.51) |
| DE.BW | 21.83 (2.29) | 15.25 (1.77) | 22.38 (3.57) | 15.57 (1.77) |
| DE.BY | 20.15 (3.01) | 15.27 (1.92) | 18.89 (4.06) | 15.41 (1.93) |
| DE.HB | 24.31 (3.1) | 25.99 (2.98) | 141.83 (104.28) | 24.87 (2.77) |
| DE.HE | 20.83 (3.77) | 15.04 (1.46) | 19.91 (4.37) | 14.99 (1.52) |
| DE.HH | 21.95 (2.73) | 21.22 (4.99) | 40.2 (10.86) | 21.54 (5.03) |
| DE.MV | 29.35 (4.57) | 28.87 (6.62) | 42.39 (8.83) | 28.61 (6.56) |
| DE.NI | 20.44 (3.45) | 15.78 (2.3) | 25.19 (4.58) | 15.93 (2.3) |
| DE.NW | 21.91 (3.27) | 12.3 (1.88) | 14.62 (2.86) | 12.53 (1.89) |
| DE.RP | 21.1 (3.5) | 15.89 (2.28) | 41.27 (16.28) | 16.12 (2.29) |
| DE.SH | 23.82 (4.2) | 19.09 (2.76) | 34.29 (8.59) | 18.41 (2.79) |
| DE.SL | 40.21 (6.97) | 35.86 (11.74) | 91.74 (28.2) | 35.52 (9.8) |
| DE.SN | 25.92 (3.26) | 19.88 (2.78) | 34.07 (6.84) | 18.98 (2.59) |
| DE.ST | 26.53 (3.32) | 21.03 (3.58) | 34.59 (8.05) | 19.55 (2.85) |
| DE.TH | 19.38 (2.62) | 15.75 (1.33) | 31.67 (6.03) | 15.42 (1.29) |

**Table S3.3.2:** Comparison of Dynamic Selection and Dynamic Stacking with and without the inclusion of metadata for COVID-19 Hospitalization in Germany. The performances are given as the mean MAPE and its standard error in parentheses. The regions are given in HASC Codes (http://www.statoids.com/ihasc.html).

| Geography | Selection | Stacking | Sel. Meta | Stack. Meta |
|---|---|---|---|---|
| DE | 27.27 (4.57) | 21.99 (2.4) | 33.68 (5.27) | 29.49 (3.86) |
| DE.BB | 32.73 (12.23) | 17.97 (2.98) | 37.21 (12.16) | 20.63 (3.09) |
| DE.BE | 39.17 (13.46) | 22.57 (5.5) | 49.69 (13.77) | 19.57 (2.23) |
| DE.BW | 30.33 (4.94) | 23.97 (3.75) | 34.81 (5.15) | 23.67 (3.04) |
| DE.BY | 35.18 (5.7) | 25.0 (2.9) | 39.83 (5.95) | 23.03 (2.88) |
| DE.HB | 42.29 (21.99) | 40.06 (22.01) | 44.38 (21.93) | 40.03 (22.04) |
| DE.HE | 26.28 (4.66) | 21.67 (2.61) | 31.86 (5.13) | 21.78 (2.91) |
| DE.HH | 33.51 (9.38) | 18.01 (2.29) | 34.83 (9.31) | 15.59 (1.46) |
| DE.MV | 36.69 (13.73) | 16.52 (1.86) | 34.66 (13.73) | 17.08 (1.93) |
| DE.NI | 26.95 (4.24) | 24.35 (2.86) | 30.24 (4.73) | 24.67 (2.62) |
| DE.NW | 24.14 (3.5) | 23.02 (2.8) | 32.09 (5.09) | 21.7 (2.83) |
| DE.RP | 37.63 (10.46) | 23.05 (3.23) | 43.44 (10.56) | 21.37 (2.31) |
| DE.SH | 35.71 (11.34) | 19.36 (2.42) | 37.15 (11.3) | 18.99 (2.31) |
| DE.SL | 52.87 (19.98) | 29.35 (7.34) | 58.06 (19.75) | 25.66 (3.91) |
| DE.SN | 35.55 (5.54) | 25.23 (3.19) | 40.86 (7.02) | 25.74 (3.29) |
| DE.ST | 31.8 (9.82) | 18.1 (2.58) | 38.86 (10.8) | 16.18 (1.69) |
| DE.TH | 32.63 (8.87) | 17.47 (1.95) | 36.91 (8.91) | 18.01 (2.02) |

**Table S3.3.1:** Comparison of Dynamic Selection and Dynamic Stacking with and without the inclusion of metadata for COVID-19 Deaths in Germany. The performances are given as the mean MAPE and its standard error in parentheses. The regions are given in HASC Codes (http://www.statoids.com/ihasc.html).
