## Supplementary figures and images for "A dynamic ensemble model for short-term forecasting in pandemic situations"

### S2_Figure.tiff

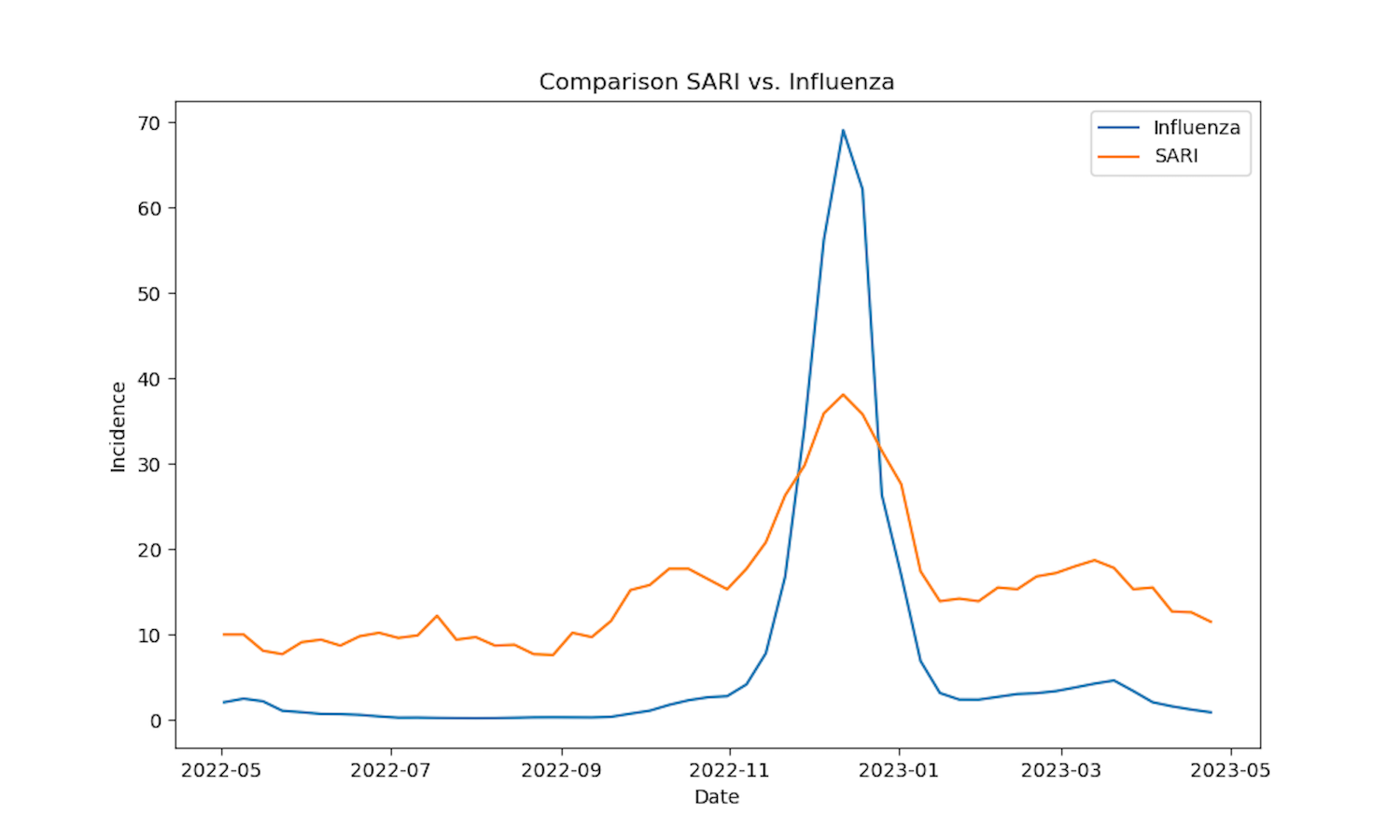
